# Systematic review and meta-analysis of COVID-19 vaccines safety, tolerability, and efficacy among HIV-infected patients

**DOI:** 10.1101/2022.01.11.22269049

**Authors:** Jacques L Tamuzi, Ley M Muyaya, Amal Mitra, Peter S Nyasulu

## Abstract

**Objective:** To conduct a comprehensive systematic review and meta-analysis of all recommended SARS-CoV-2 (Severe Acute Respiratory Syndrome Coronavirus 2) vaccines in people living with HIV (PLWH), as well as an overview of the safety, tolerability, and efficacy of the vaccines in PLWH.

**Methods:** We searched six databases, Cochrane Central Register of Controlled Trials (CENTRAL), PubMed, Medline, Medrxiv, Global research on COVID-19 database, and Google Scholar for studies investigating the effects of SARS-CoV-2 vaccines on PLWH. Results of the association were summarised by SARS-CoV IgG seroconversion and level, vaccines efficacy and tolerability. A meta-analysis was performed for studies, using random-effects model and a pooled RR with 95% CI was reported.

**Results:** Twenty-three of the 1052 studies screened met the inclusion criteria. The review included 28, 246 participants among whom 79.55% (22,469/28, 246) were PLWH with median CD4 ≥ 200 cells/µL. The pooled estimate of SARS-CoV-2 IgG seroconversion and positive neutralizing antibodies after the second vaccination dose between PLWH vs HIV negative were RR 0.95 (95%CI: 0.92 – 0.99, P = 0.006) and 0.88 (95%CI: 0.82- 0.95, P = 0.0007), respectively. The mean difference of IgG antibodies level (BAU/ml) was found higher in mRNA vaccines MD -1444.97 (95%CI: -1871.39, -1018.55). PLWH with CD4 less than 500 cells/ µl had 15% risk reduction of neutralizing antibodies response compared to those with CD4 ≥ 500 cells/µl (P = 0.003). The SARS-CoV-2 vaccine effectiveness was 65% (95%CI: 56%-72%, P <0.001) among vaccinated compared to unvaccinated PLWH. PLWH with CD4 count <350 cells/µl had lower vaccine effectiveness compared to CD4 count ≥ 350 cells/µl with 59% vs 72%, respectively. Vaccine tolerability was the same between PLWH and HIV negatives.

**Conclusion:** According to our findings, PLWH with CD4 ≥ 200 cells/µL had lower immunogenicity and antigenicity in COVID-19 vaccines than HIV negatives. Additional doses of SARS-CoV- 2 vaccination are needful in PLWH.

## 1. Background

Nearly 37.6 million [30.2 million–45.0 million] people worldwide are infected with Human Immunodeficiency Virus (HIV) [1], with Sub-Saharan Africa accounting for nearly 71% [2]. HIV and the Severe Acute Respiratory Syndrome Coronavirus 2 (SARS-CoV-2) are the main syndemics of the twenty-first century. A recent study of over 15,000 cases of COVID-19 in people living with HIV carried out by WHO revealed that PLWH were 13% more likely to be admitted to hospital with severe or critical COVID-19 after controlling for age, gender, and comorbidities [3]. Furthermore, PLWH were more likely to die after admission to hospital with COVID-19 with a 30% increased risk of death independent of age, gender, severity at presentation, and co-morbidities [3] and diabetes, high blood pressure, male sex, or over 75 years old were each associated with an increased risk of death [4]. Another study including 22 studies and 20,982,498 participants revealed that PLWH had a significantly higher risk of SARS-CoV-2 infection than HIV-negative individuals [5]. These multi-morbidities are driven by residual inflammation on ART and ongoing immune dysregulation [6] that could influence COVID-19 disease severity, the durability of protective antiviral responses, and responsiveness to vaccination [7–9]. Cellular immune deficiency and a low current or nadir CD4 cell count have been identified as potential risk factors for severe SARS-CoV-2 infection in PLWH, irrespective of HIV virological suppression [10]. Conversely, persistent inflammation or lower CD4-to-CD8 cell ratios among PLWH than among those without HIV could increase susceptibility to viral infection [11, 12]. With emerging information on PLWH with COVID-19 disease, a more pronounced immunodeficiency, defined as a current CD4 count <350/µL and a low CD4 nadir, has been associated with an increased risk for severe COVID-19 and mortality [7]. Furthermore, HIV infection is associated with chronic inflammation and immune activation, which can accelerate the aging process of the immune system, rendering it less responsive [13]. With daily antiretroviral therapy (ART), significant immune dysfunction in PLWH can be reversed and prevented [14–16]. Successful ART results in undetectable plasma HIV viraemia and a return to normal CD4+ T cell counts [16, 17]. Despite ART, HIV-associated immune pathology is not completely reversed, and many patients experience persistent T cell activation and exhaustion for years [10, 16, 18, 19]. It is therefore likely that HIV-1 infection impairs the SARS-CoV-2 immune response [20].

Despite limited data, available information suggests current WHO-recommended COVID-19 vaccines are safe for people living with HIV [21]. Several SARS-CoV-2 vaccines have proved safe and efficacious in large clinical trials, and studies are ongoing to fully characterize their safety, tolerability, immunogenicity, and efficacy. Although there are limited data in people with PLWH, mRNA, adenovirus-vectored DNA, subunit (protein), and inactivated vaccines raise no safety concerns that are specific to people with HIV [22]. Apart from this, the event of SARS-CoV-2 vaccination programs need emphasizes in PLWH. The existing data are still controversial, and gaps remain about the SARS-CoV-2 vaccines efficacy, safety, and tolerability in PLWH. Therefore, there is an urgent need to conduct a comprehensive systematic review and meta-analysis that include all vaccination against SARS-CoV-2 in PLWH and provide an overview of safety, tolerability, and efficacy, describe the immune responses of SARS-CoV-2 vaccines in PLWH. Such knowledge is critical for future COVID-19 prevention in PLWH during the pandemic, as well as informing vaccination strategies in this specific group.

## 2. Methods

### 2.1 Overview

This study followed the Preferred Reporting Items for Systematic Reviews and Meta-Analyses (PRISMA) guidelines [23]. The review protocol was registered in the International Prospective Register of Systematic Reviews (review registration number: CRD42021228863) [24].

### 2.2 Eligibility criteria

Randomized trials, open-label and non-randomized, cohort study, case-control study, and a cross-sectional study comparing different SARS-CoV-2 vaccines among which nucleic acid vaccines among (mRNA, DNA vaccines, and other popular vaccine forms), vector vaccines, inactivated vaccines, and live-attenuated vaccines between PLWH and HIV negative people, or between PLWH with different CD4 count and/or interventions.

We included adults ≥ 18 years of age with HIV, with or without a history of SARS-CoV-2 infection. All the participants included in the intervention group had proof of approved PLWH on highly active antiretroviral therapy (HAART). Additionally, PLWH with a history of SARS-CoV-2 infection was confirmed by a previous SARS-CoV-2 PCR. The non-intervention group included HIV-negative subjects and/or HIV-infected (not vaccinated or vaccinated with different SARS-CoV-2 vaccines, different from the intervention group).

### 2.3. Electronic searches

We searched eligible studies including SARS-CoV-2 vaccines in PLWH, published in English from January 2020 to November 2021 in different databases, Cochrane Central Register of Controlled Trials (CENTRAL), PubMed, Medline, Medrxiv (https://www.medrxiv.org), Global research on COVID-19 database (https://www.who.int/emergencies/diseases/novel-coronavirus-2019/global-research-on-novel-coronavirus-2019-ncov) and Google Scholar. We also searched studies on Medical Literature Analysis and Retrieval System Online, WHO International Clinical Trials Registry Platform (http://www.who.int/ictrp/en/), Global Alliance for Vaccine and Immunisation, Coronavirus Clinical Studies or COVID-19 Prevention Network (https://www.coronaviruspreventionnetwork.org/), ClinicalTrials.gov (https://www.ClinicalTrials.gov/), Conference on Retroviruses and Opportunistic Infections(CROI), HIV i-base website and International AIDS Society (IAS).

The following search strategy was conducted in PUBMED and MEDLINE: “COVID- 19”[Mesh:NoExp]) OR “SARS-CoV-2”[Majr:NoExp]) AND “HIV”[Mesh]) OR “Immunosuppression”[Majr]) OR “Immunocompromised Host”[Majr]) AND “Vaccines”[Majr])) OR “Vaccination”[Mesh]) OR “Immunization”[Majr]) OR “Immunogenicity, Vaccine”[Majr]) AND “Safety”[Majr:NoExp]) OR “Treatment Outcome”[Majr]. The following keywords were used to search studies on other databases and conference websites: “COVID-19 vaccines, SARS-CoV-2 vaccines, HIV, HIV-infected patients, immune-depression, immunocompromised, efficacy, and tolerability”. (SARS-CoV-2 vaccines OR COVID-19 vaccines) and (HIV-infected people OR HIV OR immunocompromised OR immunosuppression) and (SARS-CoV-2-specific antibodies OR SARS-CoV-2 spike antibody responses).

### 2.4. Study selection

Duplicate studies were deleted electronically using EndNote’s “Find Duplicates” tool, and all studies collected from electronic databases using the search method were imported into EndNote V.X9.5. The studies were manually reviewed again to identify and remove any additional duplicates. Two authors (JLT and LMM) choose papers that are unrelated to one another. For any titles and abstracts that appear to fit the inclusion requirements or where there is any doubt, complete articles were collected. Inappropriate population, improper intervention, improper comparison, inappropriate outcome(s), inappropriate study design, and others were excluded. Following selection, the two authors (JLT and LMM) discussed their choices and reached an agreement on any disparities. If JLT and LMM authors were unable to achieve an agreement, PSN was consulted. Cohen’s kappa (κ) statistics were used to calculate the overall agreement rate before discrepant items are corrected. Following the resolution of conflicts, the overall precision of searches was calculated by dividing the number of studies included by the total number of studies screened after duplicates were removed. The degree of agreement between the two independent data extractors (JLT and LMM) was computed using kappa statistics to indicate the difference between observed and expected agreements at random or by chance only. The following was how the Kappa values were interpreted: (1) less than 0 equals less than chance agreement, (2) 0.01– 0.20: small agreement, (3) 0.21–0.40: good agreement, (4) 0.41–0.60: moderate agreement, (5) 0.61–0.80: significant agreement, and (6) 0.8–0.99: practically perfect agreement [25].

### 2.5. Quality assessment

Two reviewers (JLT and LMM) independently assessed the risk of bias in the included papers. Any disagreement between the two reviewers about the risk of bias was settled by a conversation with the third reviewer (PSN). For randomized controlled trials, the Cochrane risk bias assessment tool was used [26]. Random sequence creation, concealment of allocations, blinding of participants and staff, blinding of outcome assessments, insufficient data on outcomes, and selective reporting were all part of the risk assessment [26]. A bias risk assessment tool in non-randomized intervention studies (ROBINS-I) was used to assess the risk of bias in included non-randomized studies, including bias due to confounding (pre-intervention), bias in the selection of study participants (pre-intervention), and bias in the classification of interventions (intervention), bias due to deviations from the intended intervention (post-intervention), bias due to missing data (post-intervention), bias in the measurement of outcomes (post-intervention), bias in the selection of reported outcomes (post-intervention) and overall bias risk [27]. We assessed the risk of bias as the low, moderate, serious, critical risk of bias, and no information [27].

### 2.6. Data abstraction

Data were extracted by two independent reviewers (JLT and LMM) using a standardized data abstraction form, developed according to the sequence of variables required from the primary studies. Disagreements in data abstraction between the reviewers were resolved by a third independent reviewer (PSN). All data were extracted into an Excel worksheet template. Data were extracted on the following: author’s first name, publication date and location (country in which the research was conducted), methods (study design, study setting, sample size, total study duration, withdrawal, and date of study), participants (sample size, gender, mean age, median and ranges CD4 count, HIV viral load (suppressed or not suppressed), inclusion and exclusion criteria), interventions (COVID-19 nucleic acid vaccines, vector vaccines, inactivated vaccines and live attenuated vaccines), and comparisons included PLWH with placebo, no intervention or different vaccines compared to the intervention group. We also included as comparisons HIV-negative with placebo, no intervention, and same vaccine as the intervention group.

The primary and secondary outcomes were specified and collected, and time points were reported. We included primary outcomes among which seroconversion for SARS-CoV-2 neutralizing antibody after the first and the second doses of the vaccines, IgG antibody level after the first and second vaccines doses, anti-SARS-CoV-2 neutralizing antibodies, SARS-CoV-2 vaccines effectiveness, local and systematic reactions after the first and second doses. The second outcomes included PLWH subgroups based on ≥350 cells/µl; and <500 vs ≥500 cells/µl.

### 2.7. Interventions

The interventions included any type of SARS-CoV-2 vaccine regardless of dose (1, 2, or 3 doses), administration routes, and duration. Based on the mechanism of action of SARS-CoV-2 vaccines, we grouped them as inactivated or weakened virus vaccines, protein-based vaccines, viral vector vaccines, RNA and DNA vaccines administered as primary series of one or two doses according to vaccines protocols.

### Inactivated vaccines

Are kind of whole-cell vaccine in which the pathogenic materials of the causing pathogen or a very similar one are destroyed by chemicals (such as formaldehyde or beta-propiolactone), radiation, or heat. This demolishes the pathogen’s ability to replicate while keeping its immunization potential [28, 29]. One of the inactivated vaccines manufactured for the COVID-19 infection is CoronaVac® or Sinovac® containing the inactivated virus and aluminium as an adjuvant.

### Protein subunit vaccines

Subunit vaccines merely consist of the immunogenic proteins derived from a pathogen, which can stimulate the host’s immune system [29]. The proteins can be easily produced by recombinant DNA techniques [29, 30]. It was shown that protein subunit vaccines together with adjuvants generated a potent response against SARS-CoV-2 [29]. The vaccines have good safety profiles since there are no live components; reactogenicity and side effects are mainly related to adjuvants [31]. In the case of Novavax, neutralizing antibodies (nAbs) was 100-times higher after the second dose and 4-times higher than symptomatic outpatients after 35 days [29, 32].

### Viral vector vaccines

A viral vector is a modified virus used to deliver a target gene to the host cell. Modified virus is obtained by inserting an immunogenic part of the desired virus, which is the S protein gene for SARS-CoV2 into a safe and non-pathogenic virus as a vector [29, 33]. Subunit Vaccines targets are S proteins in the development of subunit vaccines, which would lead to inhibition of the binding of the viruses to the host angiotensin-converting enzyme 2 (ACE2) [34, 35]. There are two types of viral vector vaccines: non-replicating and replicating. The non-replicating vectors do not have genes responsible for replication, so they cannot replicate and only produce virus antigens. On the other hand, replicating vectors, multiply in the body and infect other cells, causing all cells to produce antigens [36, 37]. Viral vector vaccines include ChAdOx1 nCoV- 19, Sputnik V, Ad26.COV2-S, Ad5-nCoV, and others.

### Nucleic Acid Vaccines

are usually made of mRNA molecules encapsulated in lipid nanoparticles (LNP). LNP facilitates mRNA penetration into the host cell so that the translation process can be initiated [28, 29]. The cellular immune response is seen when the S proteins are presented to immune cells either by the MHC class I or II complex, activating both CD4^+^ and CD8^+^ cells [29]. The salient features of mRNA vaccines are: (i) Translation of mRNA occurs in the cytoplasm to avoid the risk of any genome integration. (ii) Both T cell and antibody responses are triggered. (iii) Rapid development and large-scale mass production can be achieved [31]. Two important mRNA vaccines for COVID-19 are mRNA-1273 by Moderna TX, Inc, BNT162b1 by BioNTech- Fosun Pharma-Pfizer, and LNP-nCoVsa RNA. A DNA vaccine is another type of nucleic acid vaccine. DNA vaccines are produced by inserting the gene of an antigenic protein into a bacteria-derived plasmid and delivering it to the host cells. The protein-producing machinery of the host cell will translate DNA to mRNA and finally, a protein that can stimulate the immune system.

### 2.8. Outcomes

#### 2.8.1. Primary outcomes

**IgG antibody seroconversion for SARS-CoV-2 after the first and the second doses of the vaccines:** defined as numbers or proportions of IgG seropositivity status such as ELISA specific SARS-CoV-2 spike receptor-binding domain (RBD) antibody responses among PLWH and HIV negatives.

**IgG antibody level after the first and second doses:** measured total binding antibodies against SARS-CoV-2 nucleocapsid (N) and spike receptor-binding domain (RBD). Results reported in arbitrary units (AU)/mL were converted to binding antibody units (BAU/ml).

**SARS-CoV-2 neutralizing antibodies (NAbs) after the second vaccine dose:** Antibodies that bind to Spike, a large homotrimeric glycoprotein studded across the viral surface, have been shown to neutralize SARS-CoV-2 [35, 36]. NAbs are critical for virus clearance and SARS-CoV-2 protection [37]. This outcome included numbers and proportions of PLWH vs HIV negatives with high titers of neutralizing anti-S antibodies.

**SARS-CoV-2 vaccine efficacy (%)** was defined as (1– Relative Ratio) ×100, where RR is the relative risk of Covid-19 illness including mild, moderate, severe, and critical cases confirmed by Polymerase chain reaction (PCR) among vaccinated and unvaccinated PLWH.

**Local and systematic reactions after the first and second doses:** Numbers and proportions of PLWH vs HIV negatives with local and systemic AEs.

#### 2.8.2. Secondary outcomes

The outcomes included PLWH subgroups based on positive neutralizing antibodies with 200 cells/µl and <500 vs ≥ 500 cells/µl: and vaccines effectiveness <350 vs ≥350 cells/µl. We also compared IgG antibody levels between different types of vaccines.

### 2.9. Statistical analysis

The meta-analysis was carried out with the review manager software (Revman version 5.3) [38] and complementary analyses were undertaken with ProMeta version 3 [39]. We presented data from eligible studies in evidence tables and figures (map, flow diagram, forest plots, funnel plot, and meta-regression figures). The relative risks (RR) with 95% CI of the effect size for binary outcomes and the mean difference (MD) with 95% CI were reported for continuous outcomes. In cases where continuous outcomes were reported in median (IQR), the conversion to mean (SE) was undertaken according to [40]. We converted AU/ml to BAU/ml with the conversion factor as follows: 1 BAU/mL = 0.142 × AU/mL. Pooled results from continuous outcomes have been expressed in BAU/mL [41].

Based on the type of SARS-CoV-2 vaccines (viral vector, protein subunit, mRNA, DNA, and inactivated vaccines), we built different forest plots including subgroup analysis of vaccines type for the following outcomes: RBD IgG seroconversion, IgG antibody level, neutralization antibodies responses, vaccine efficacy, SARS-CoV-2 post-vaccination, local reaction, and systemic reactions. As statistical heterogeneity was significant because of different vaccines groups, the random-effect model (Mantel-Haenszel or inverse variance methods) was adopted in this meta-analysis. Heterogeneity between the results of the primary studies was assessed using Cochran’s Q test and quantified with the I-squared statistic. A probability value less than 0.1 (p*<*0.1) was considered to suggest statistically significant heterogeneity [42]. Heterogeneity was considered low, moderate, and high when the values are <25%, 25% to 75%, and >75%, respectively [42]. In case of high heterogeneity (above 90%) after subgroup analysis, the results were reported by type of vaccines or individual studies.

We computed Egger’s regression and Begg Mazumdar’s rank correlation test to evaluate possible publication bias. We conducted a sensitivity analysis to assess the robustness of our pooled RR of RBD antibody responses after the first and the second dose of SARS-CoV-2 vaccines among PLWH and HIV-negative people. The quality of evidence was assessed for each study using Grading of Recommendations, Assessment, Development and Evaluation (GRADE) criteria [43]. The Grade was assessed through the study design and risk of bias, inconsistency, indirectness, imprecision, and publication bias [43].

## 3. Results

### 3.1. Characteristics of included studies

A total of 1122 studies were identified in the databases, of which 70 were eliminated as duplicates, 1052 by reading titles and abstracts, 65 by reading the full text, and 42 were excluded with reasons as mentioned in **Figure 1**. Twenty-three studies were selected for the analysis [16, 44–65]. The review included 28, 246 participants among whom 79.55% (22,469/28, 246) were PLWH. All studies included participants 18 years and above. All the medians’ CD4 count was≥ 200 cells/µl (Supplementary material: Table 1). Sixteen studies reported the HIV viral load, among them, six studies included PLWH with viral load < 50 copies/ml [16, 45, 49, 51, 53, 54] and nine other studies included PLWH with viral load 58.1 to 95% suppressed [44, 46, 52, 55, 56, 58, 60, 61, 63, 65] (**Supplementary material: Table 1**).

**Figure 1:**
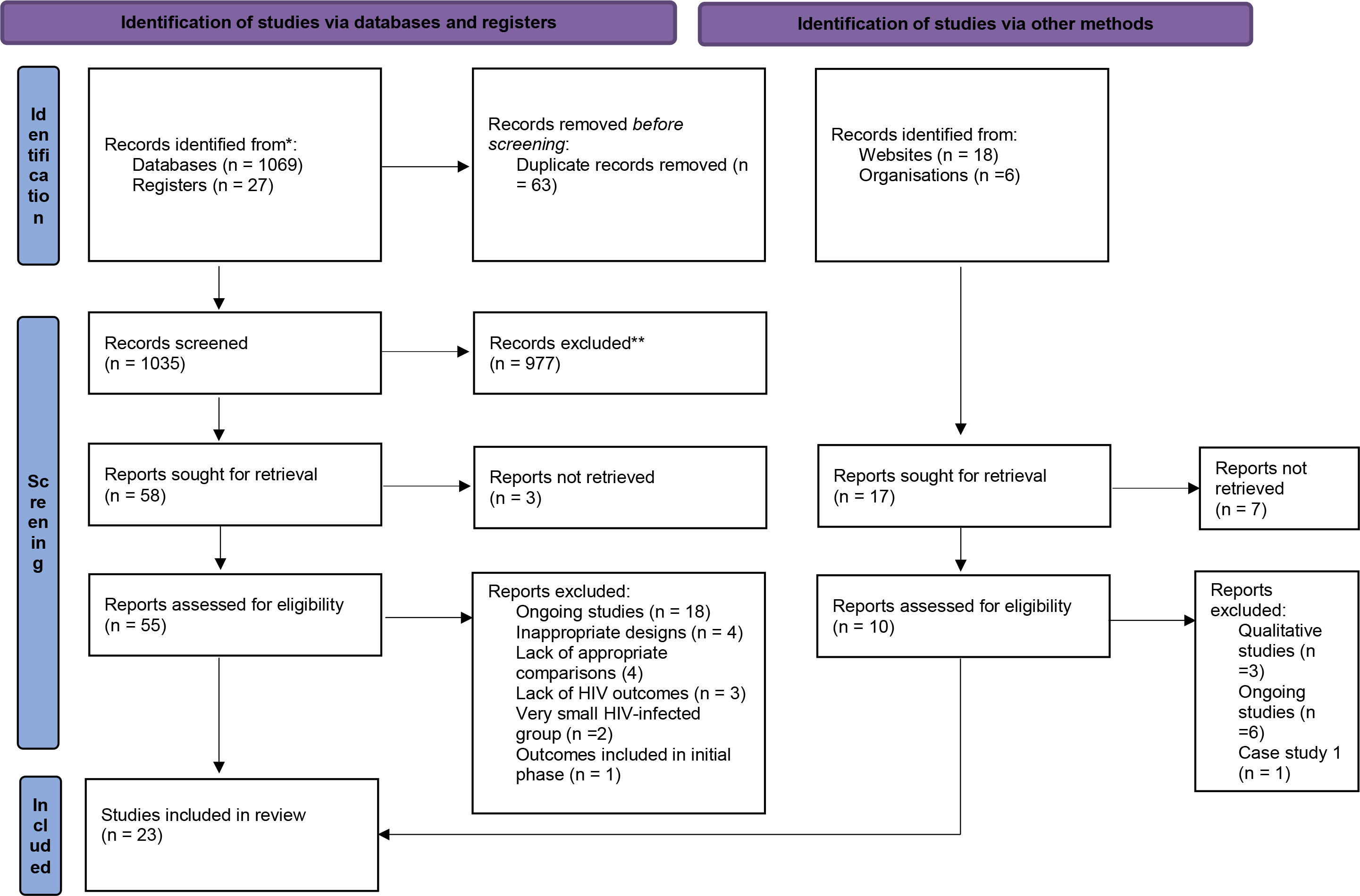
Flow diagram of COVID-19 vaccines safety, tolerability, and efficacy among HIV-infected patients

The selected articles reported data from United States of America (n = 4) [48–50, 55], China (n=5) [52, 54, 59, 61, 64], South Africa (n = 3) [45, 46, 57], United Kingdom (n = 2) [16, 53], Canada (n=2) [ 47, 51], Italia (n=1) [58], Brazil (n =1) [56], Germany (n =1) [60], Israel (n = 1) [44], Sweden (n=1) [64], Taiwan (n=1) [65] and Russia (n = 1) [62] (**Figure 2**). Ten studies used randomized control designs [16, 45, 46, 49, 50, 53, 54, 64, 64, 65], eight studies used prospective cohort designs [44, 47, 48, 51, 52, 56–58], two retrospective cohort studies [62, 63], two cross sectional studies [59, 61] and one case control study [55] (**Supplementary material: Table 1**). Eleven studies included mRNA vaccines among which BNT162b2 mRNA, Vaccine, mRNA-1273 (100 μg) administered intramuscularly (IM) as a series of two doses (0.5 μmL each), given 28 days apart [44, 47, 48, 50, 51, 53, 55, 58, 60, 63, 64], six studies included inactivated vaccines μg/0.5 mL) or Sinovac (0.5 μml) given intramuscularly [52, 54, 56, 59, 61, 64], seven studies included viral vector vaccines among which ChAdOx1 nCoV-19 with two doses (5 × 1010 vp) was given 4-6 weeks apart, Ad26.COV2.S [16, 45, 49, 51, 57, 62, 63] and two study included sub unit g (sub unit) of recombinant spike protein with 50μg of Matrix-M1 adjuvant) administered two intramuscular injections, 21 days apart and two standard doses of 15 mcg MVC-COV1901, administered 28 days apart via IM injection [46, 65] (**Supplementary material: Table 1**)

**Figure 2:**
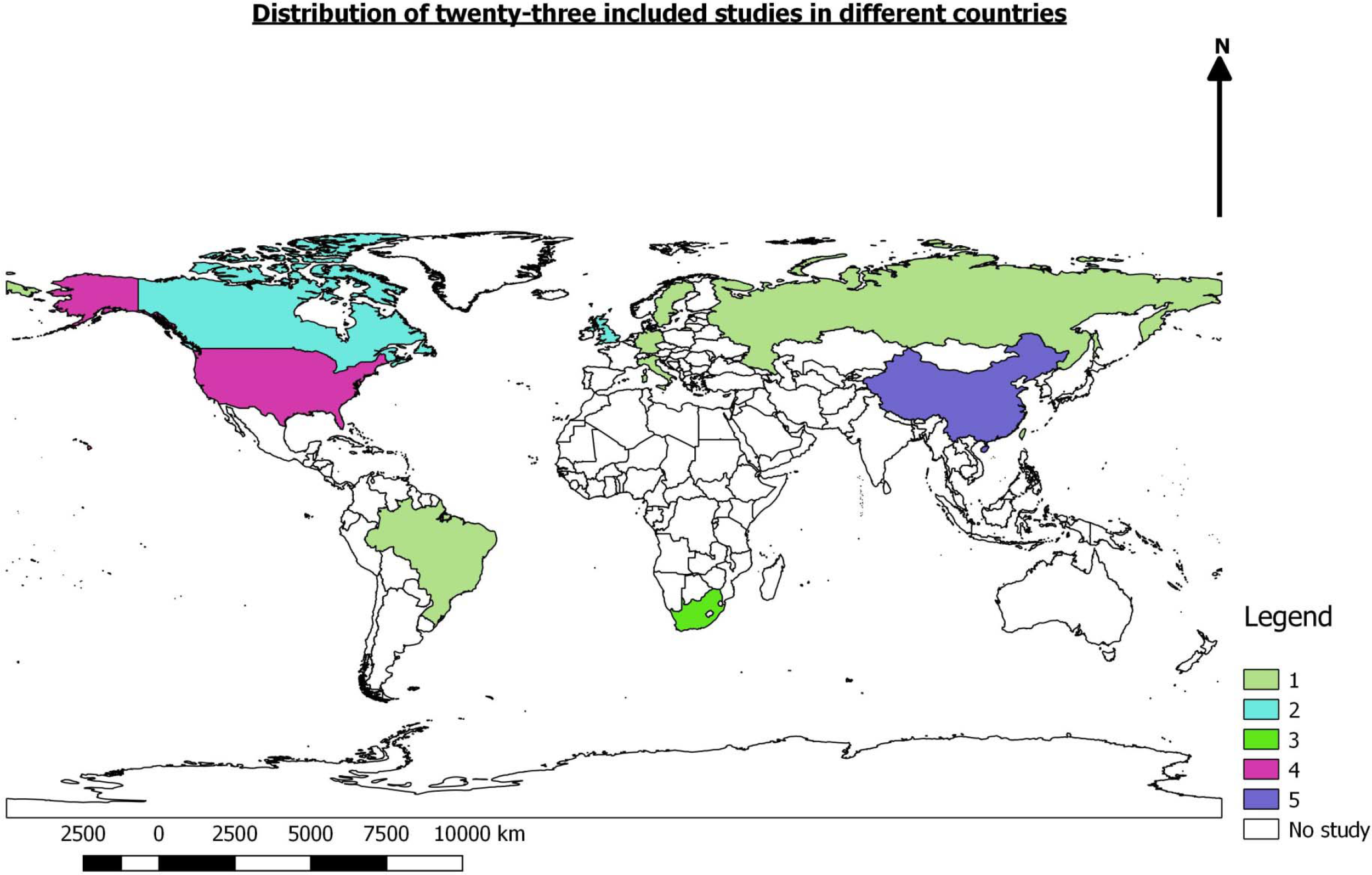
Distribution of twenty-three included studies in different countries

In terms of the bias risk analysis of the domains considered in the Cochrane Collaboration tool [25], all eight randomized control trials [16, 45, 46, 49, 50, 63–65] showed an unclear for allocation concealment bias risk (**Supplementary material: Table 2**). Four papers reported a high risk of bias for random sequence generation as well as masking of participants and professionals [16, 63–65], five trials showed a low risk of blinding of participants and personnel [45, 46, 49, 50, 65], one study reported a high risk of blinding of outcomes assessment [63], two studies reported a low risk of other bias [49, 50], and others reported a high risk. In other domains, all studies presented low bias risks (**Supplementary material: Table 2**). Regarding the risk of bias of non-randomized studies, sixteen studies were assessed with the ROBIN-I bias tool assessment [26]. Among them, seven were rated as low-moderate for the overall ROBIN-I [44, 51, 52, 55, 57, 58, 60], four studies were low–serious [53, 54, 56, 62], one study was moderate-serious [61], another study was Serious–Critical [48] and two studies were critical [47, 59] (**Supplementary material: Table 3**).

### 3.2. Meta-analysis

#### 3.2.1. IgG seroconversion after 1^st^ vaccination dose

**Figure 3** shows a comparative forest plot analysis of IgG seroconversion after the first SARS-CoV-2 vaccines across published eight studies [16, 44, 45, 49, 52, 56] looking at the RR between 935 PLWH with CD4 ≥ 300 cells/µl and 1013 HIV negative people.

**Figure 3:**
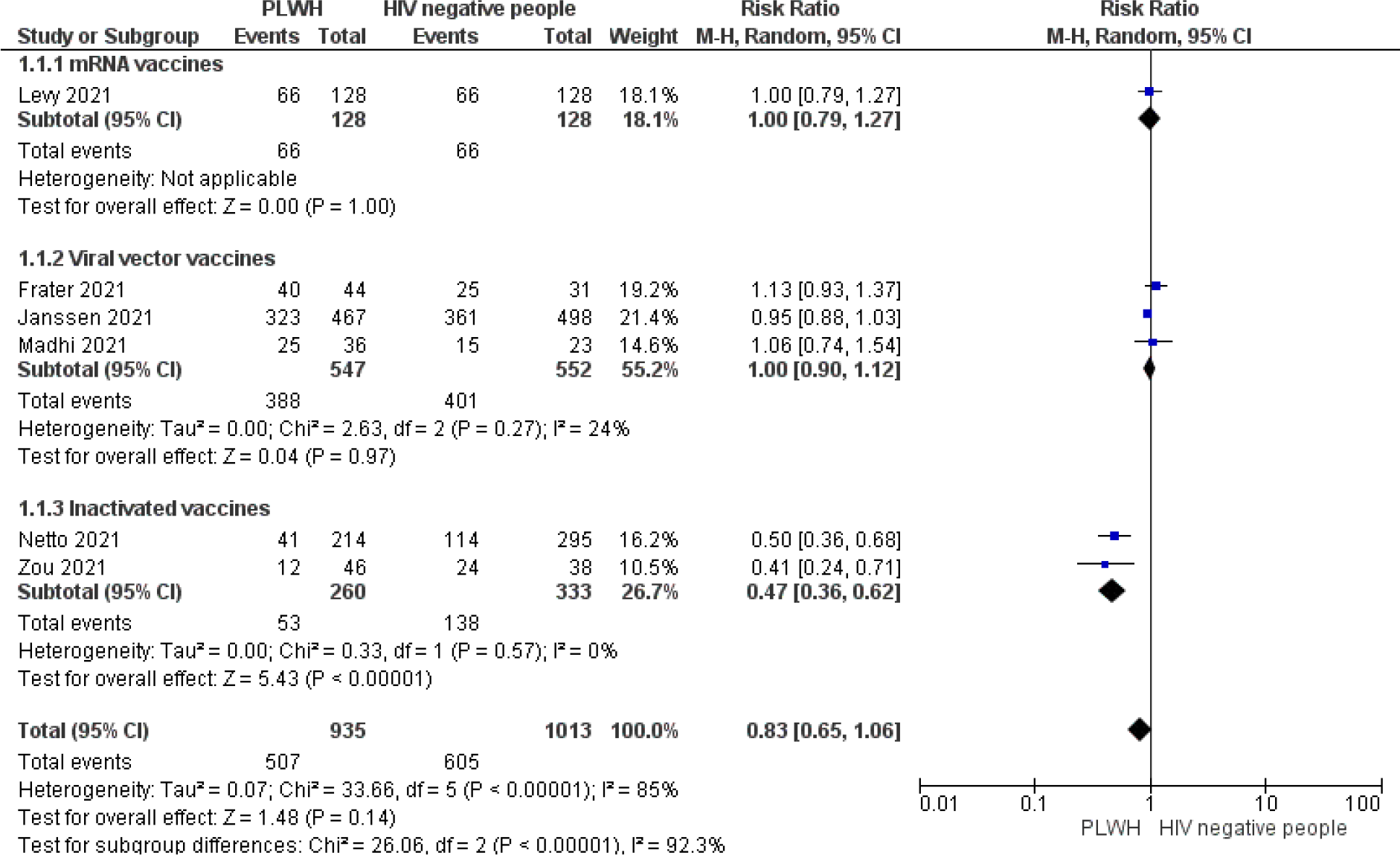
Forest plot analysis of the overall risk ratio RBD antibody responses after the first dose of SARS-CoV-2 vaccines among PLWH and HIV-negative people.

Overall, the random-effects model presented a RR of 0.83 (95%CI 0.65–1.06, 1948 participants, P = 0.14, I^2^ = 85%). The results presented by type of vaccine showed SARS-CoV-2 inactivated vaccines administered in PLWH had 53% risk reduction of IgG seroconversion compared to HIV negative people. In contrast mRNA and viral vaccines did not have different seroconversion between PLWH and HIV negative people with RR 1.00(95%CI: 0.79-1.27) and 1.00(95%CI: 0.90-1.12), respectively.

#### 3.2.2. IgG seroconversion after 2^nd^ vaccination dose

The pooled seroconversion results after SARS-CoV-2 vaccination second dose between 1885 PLWH and 7768 HIV negative people showed statistically difference between the two groups RR 0.95 (95%CI: 0.92 – 0.99, P = 0.006), with the test for subgroup differences I^2^ = 41.3% (p = 0.16) (**Figure 4**). IgG seroconversion after the second vaccination dose was not statistically different between PLWH and HIV negative people for mRNA and protein subunit vaccines with RR 0.98 (95%CI: 0.96-1.00, P = 0.06) and 0.91(95%CI: 0.42-1.98, P = 0.81), respectively. In contrast, viral vector and inactivated vaccines reported statistically lower IgG seroconversion after second vaccination dose among PLWH with RR 0.93 (95%CI: 0.88-1.00, P = 0.04) and 0.90(0.81-0.99, P = 0.03), respectively.

**Figure 4:**
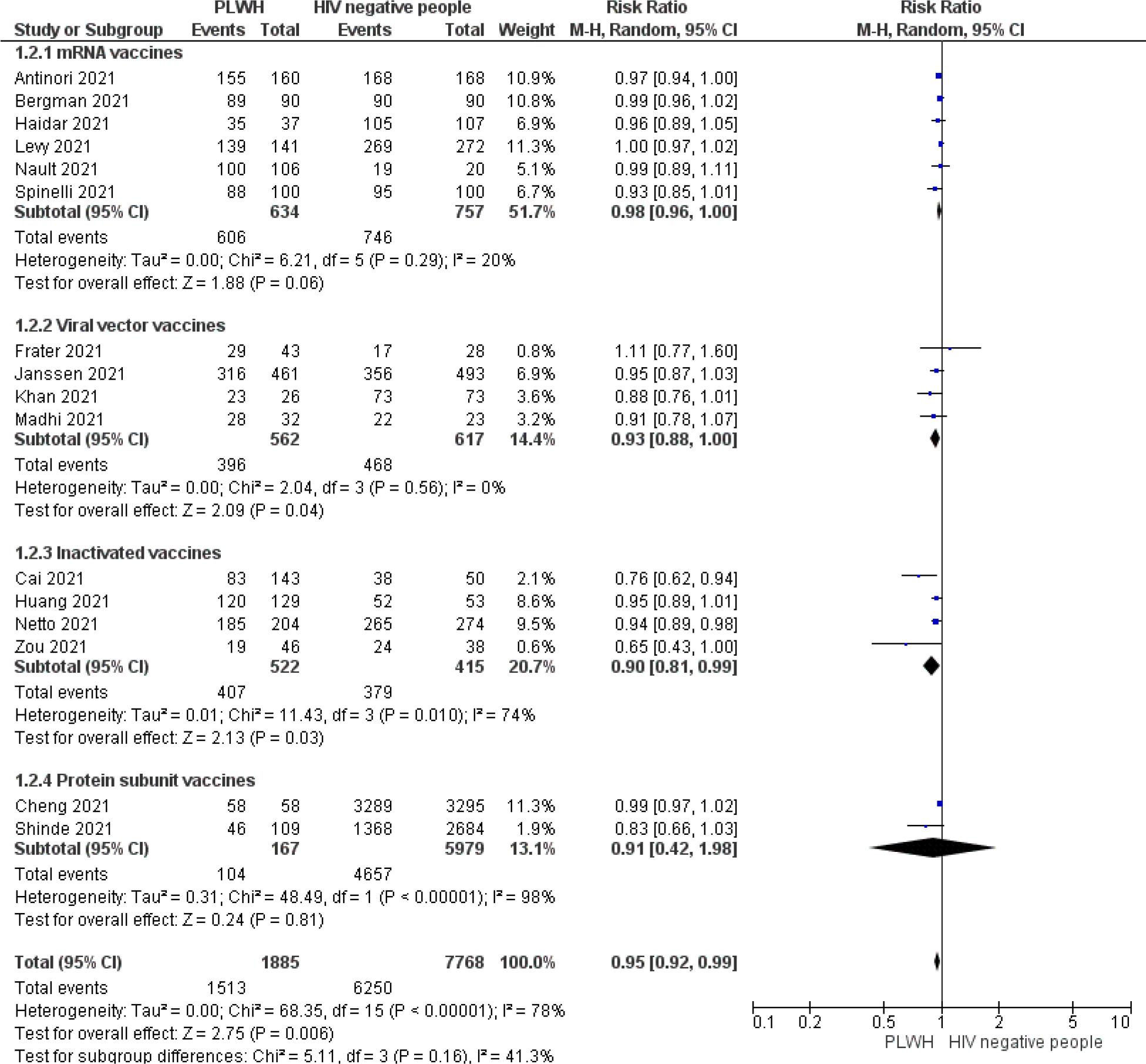
Forest plot analysis of the overall risk ratio RBD antibody responses after the second dose of SARS-CoV-2 vaccines among PLWH and HIV-negative people.

#### 3.2.3. IgG antibody level after the first dose of SARS-CoV-2 vaccines (BAU/ml)

**Figure 5** reported IgG antibodies level after the first dose of SARS-CoV-2 vaccines. The pooled result including eight studies [44, 45, 51–54, 56, 60] has shown that the mean difference (MD) IgG antibody level (BAU/ml) did not differ between 643 PLWH and 762 HIV negatives after the first dose of SARS-CoV-2 vaccines MD-0.02 (95%CI: -0.09- 0.05, P = 0.58). The test of heterogeneity between studies was high (I^2^ = 83%). However, the test for subgroup difference was not statistically significant (I^2^ = 33.6%, P = 0.21).

**Figure 5:**
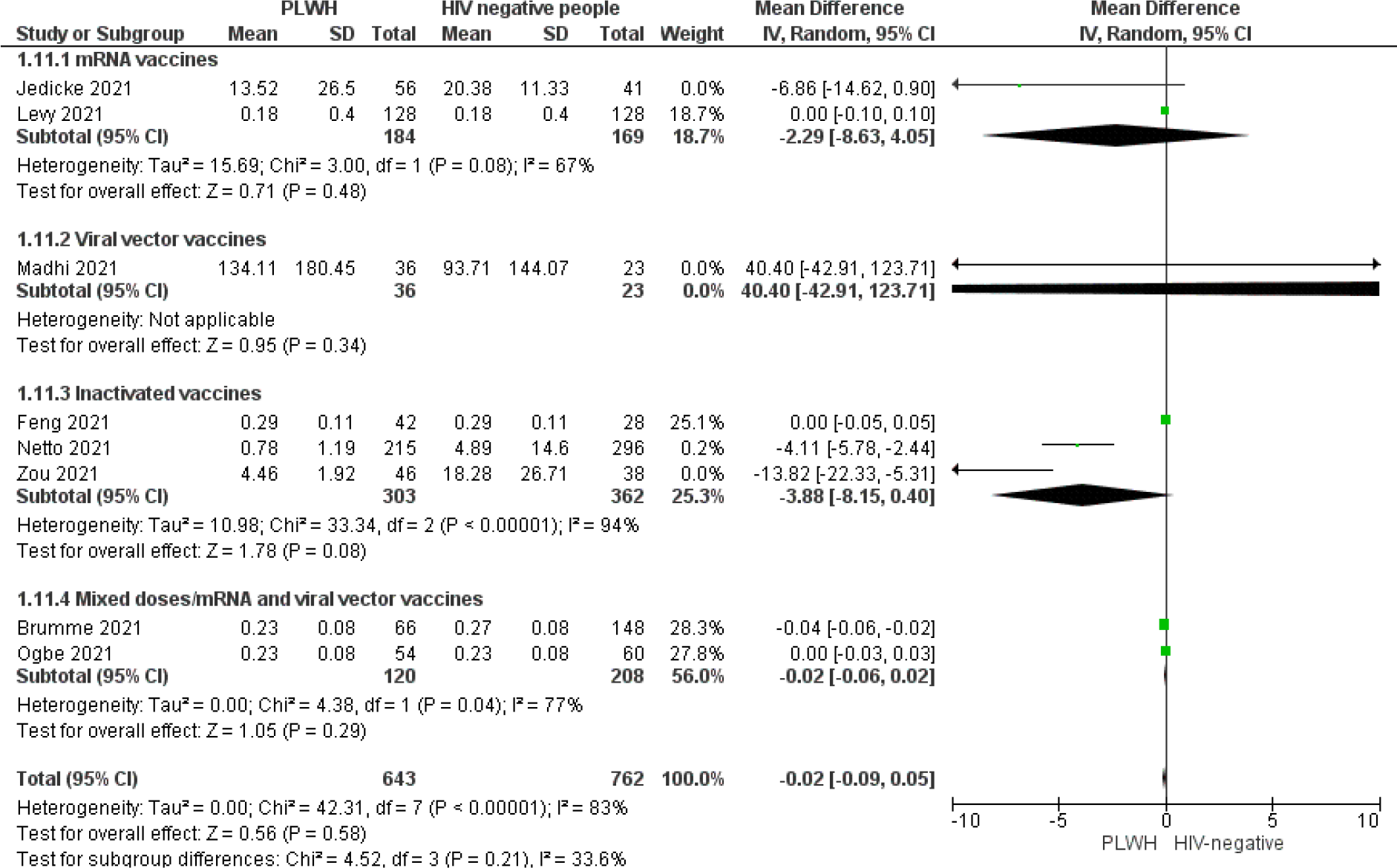
Forest plot of mean difference IgG antibody level (BAU/ml) after the second dose of SARS-CoV-2 vaccines

#### 3.2.4. IgG antibody level (BAU/ml) after the second dose of SARS-CoV-2 vaccines

Twelve studies reported IgG antibody levels after the second SARS-CoV-2 dose (**Figure 6**). As heterogeneity was high between included studies and /or vaccines type, the results were reported narratively. Eight studies out of eleven showed a statistically significant decrease of the mean difference IgG antibody level after the second SARS-CoV-2 dose (BAU/ml) among PLWH compared to HIV-negative people. The highest and lowest statistically significant mean difference IgG antibody level (BAU/ml) after the second SARS-CoV-2 dose were found in mRNA vaccines MD -1444.97 (95%CI: - 1871.39, -1018.55) [58] and MD -0.80(95%CI: -0.90, -0.70).

**Figure 6:**
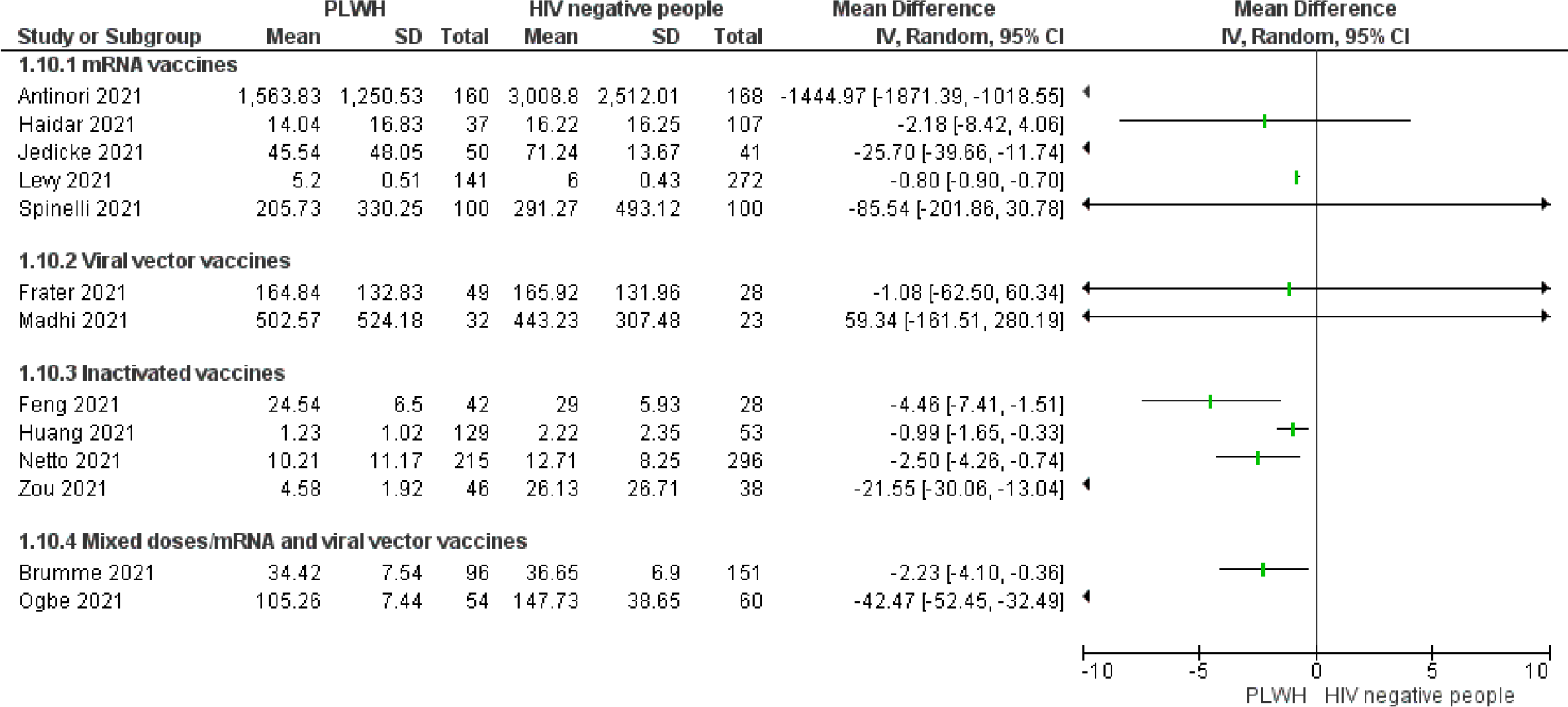
Forest plot of mean difference IgG antibody level (BAU/ml) after the second of SARS-CoV-2 vaccines

#### 3.2.5. SARS-CoV-2 vaccines effectiveness

**Figure 7** described the RR of COVID-19 vaccines effectiveness among 20,314 vaccinated and unvaccinated PLWH. The pooled results included four studies [46, 49, 50, 62] showed that that vaccine effectiveness was 65% (95%CI: 56%-72%, P <0.001) among vaccinated compared to unvaccinated PLWH, with the test for subgroup differences I^2^ = 0% (P = 0.47).

**Figure 7:**
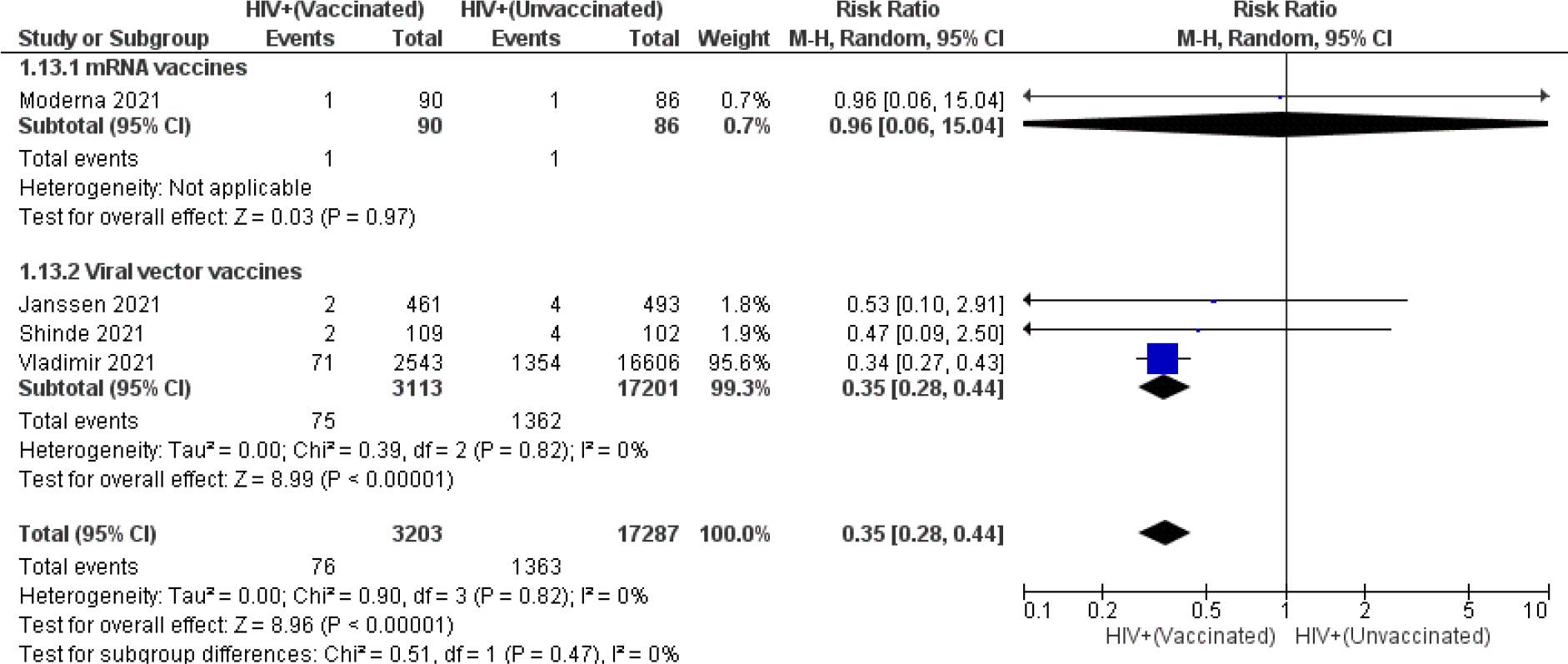
Meta-analysis of SARS-CoV-2 vaccines effectiveness: vaccinated vs unvaccinated PLWH.

#### 3.2.6. Vaccine’s effectiveness based on CD4 count

The pooled SARS-COV-2 vaccines effectiveness of 13,547 vaccinated and unvaccinated PLWH with CD4 count <350 vs ≥ 350 cells/µl was 69 % (95%CI: 58%- 78%, P < 0.001) (**Figure 8**). However, PLWH with CD4 count <350 cells/µl had lower vaccine effectiveness compared to CD4 count ≥ 350 cells/µl with 59% vs 72%, respectively. The test for subgroup differences was not statistically different between the two groups (I^2^ = 23.2%, P = 0.25) (**Figure 8**).

**Figure 8:**
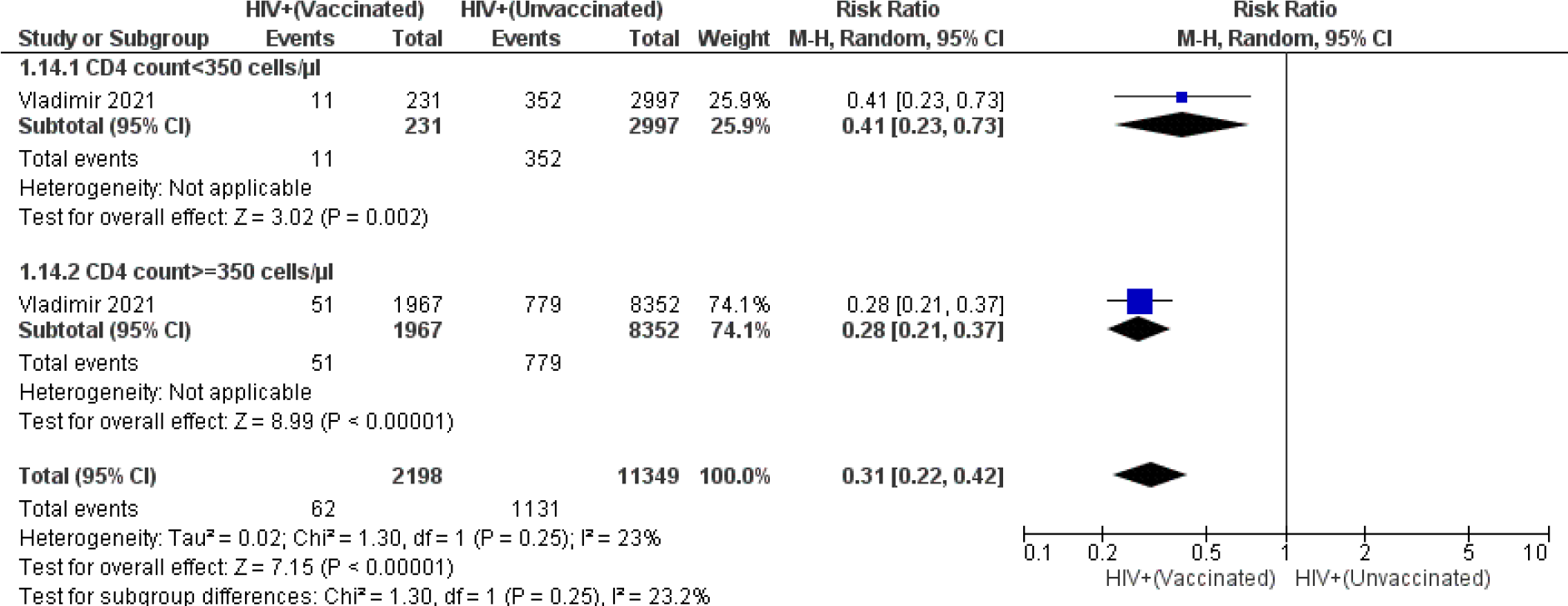
Meta-analysis of SARS-CoV-2 vaccines effectiveness based on CD4 count

#### 3.2.7. SARS-CoV-2 vaccines neutralizing antibodies response after the second dose

**Figure 9** reported the pooled neutralizing antibodies response after the second dose of SARS-CoV-2 vaccines between 769 PLWH and 613 HIV-negative people. PLWH had 12% risk reduction of neutralizing antibodies response compared to HIV negative people with RR 0.88 (95%CI 0.82-0.95, 1,059 participants, P = 0.0007). mRNA vaccines had statistically significant lower neutralizing antibodies response RR 0.89 (95%CI 0.84- 0.93, P < 0.0001). In contrast, neutralizing antibodies response between PLWH and HIV negative people was not statistically significant with inactivated vaccines, RR 0.88 (95%CI 0.77-1.02, P = 0.10).

**Figure 9:**
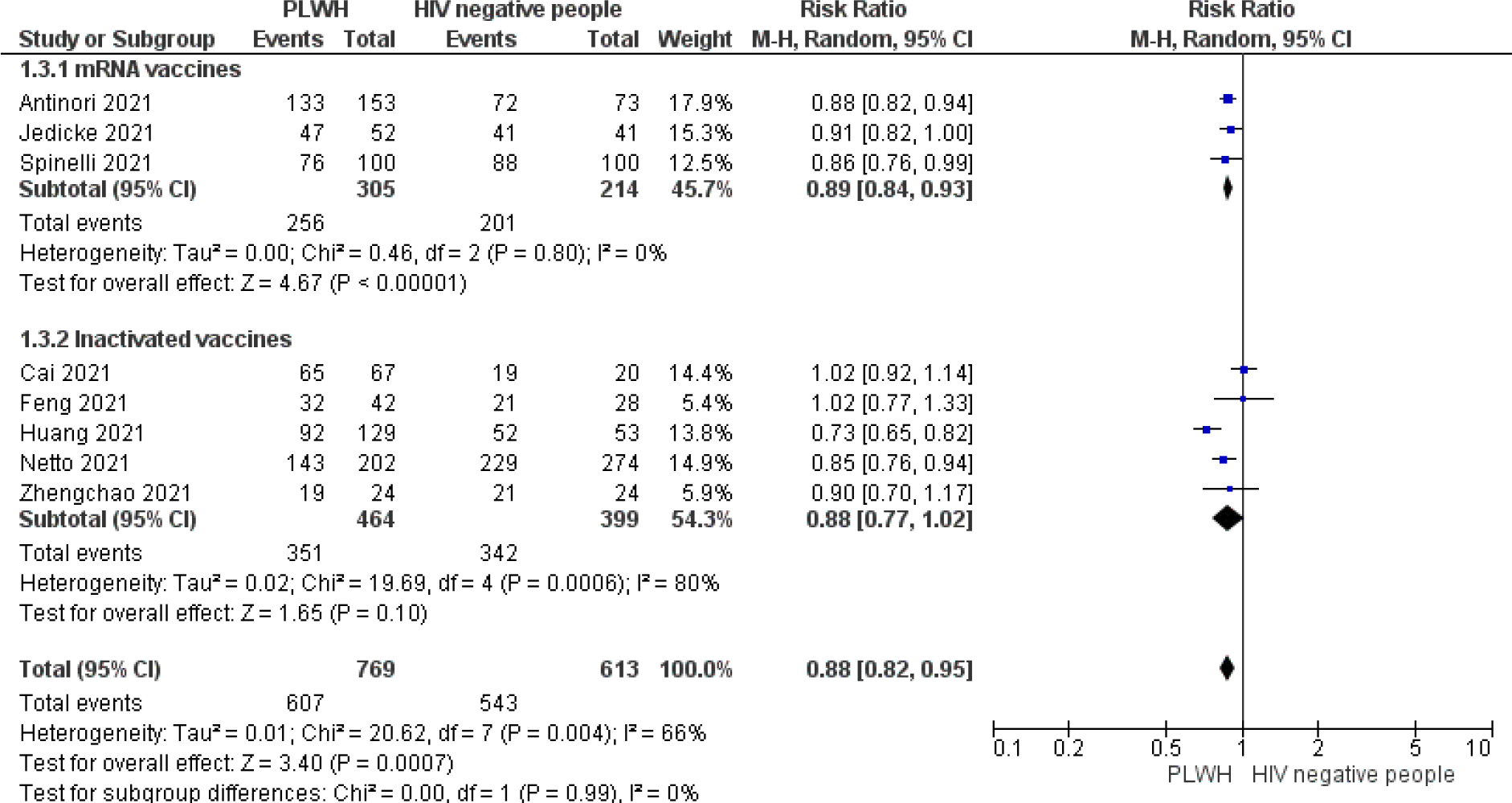
Pooled results of SARS-CoV-2 vaccines neutralizing antibodies response after the second dose

#### 3.2.8. Any local reaction after the first SARS-CoV-2 vaccines dose

The meta-analysis including five studies with local reactions after the first SARS-CoV-2 vaccines dose did not show statistically significant between PLWH and HIV negative people with RR 0.98 (95%CI: 0.71 – 1.36, 1,688 participants, P = 0.91), with the test for subgroup differences I^2^ = 80.9.0% (p = 0.001). However, Levy and al. showed significant local reactions among HIV negative people RR 1.59 (95%CI: 1.22-2.07, P = 0.0006) (**Figure 10**).

**Figure 10:**
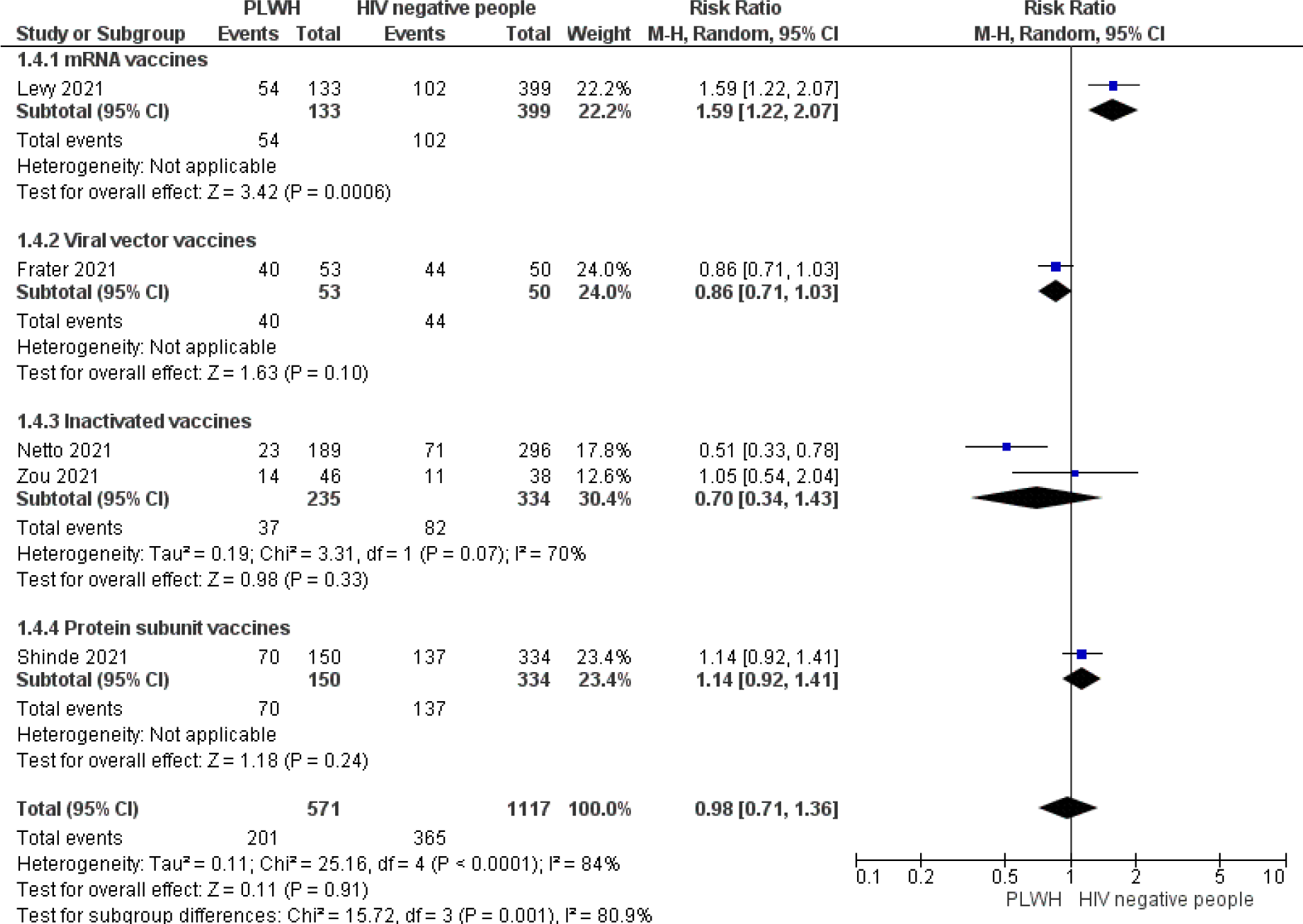
Forest plot analysis compared the overall risk ratio of local reactions after the first dose of SARS-CoV-2 vaccines among PLWH and HIV negatives.

**Figure 11:**
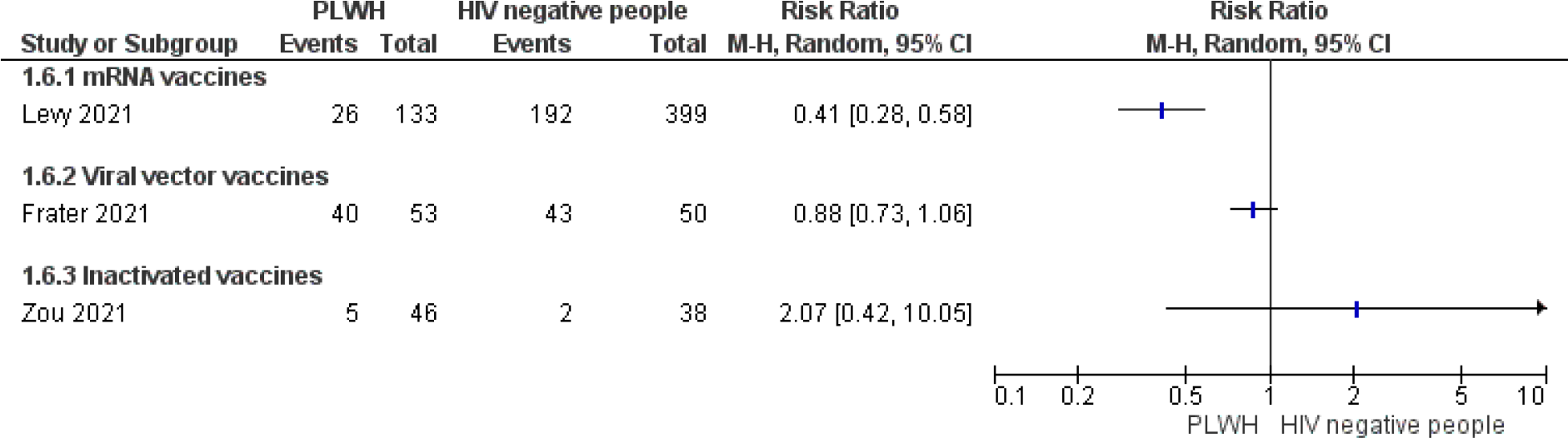
Forest plot of systemic reactions after the first dose of SARS-CoV-2 vaccines: PLWH vs HIV negative participants

#### 3.2.9. Any systemic reaction after the first SARS-CoV-2 vaccines dose

Heterogeneity between protein subunit, mRNA, and the viral vector vaccines was high in the meta-analysis of systemic reactions after the first SARS-CoV-2 vaccines dose, therefore we reported the results separately. mRNA vaccine reported a lower risk of systemic reactions after the first SARS-CoV-2 vaccine among PLWH compared HIV negative people RR 0.41 (95%CI: 0.28 – 0.58). In contrast, both inactivated and viral vector vaccines reported the same risk of systemic reaction after the first dose of SARS-CoV-2 vaccines among PLWH and HIV-negative participants.

#### 3.2.10. Any local reaction after the second SARS-CoV-2 vaccines dose

The estimated RR of local reactions after the second SARS-CoV-2 vaccines dose between 838 PLWH and 4, 533 HIV-negative people was not statistically different RR 0.97 (0.82 – 1.14, P = 0.06), with the test for subgroup differences I^2^ = 15.9% (**Figure 12**).

**Figure 12:**
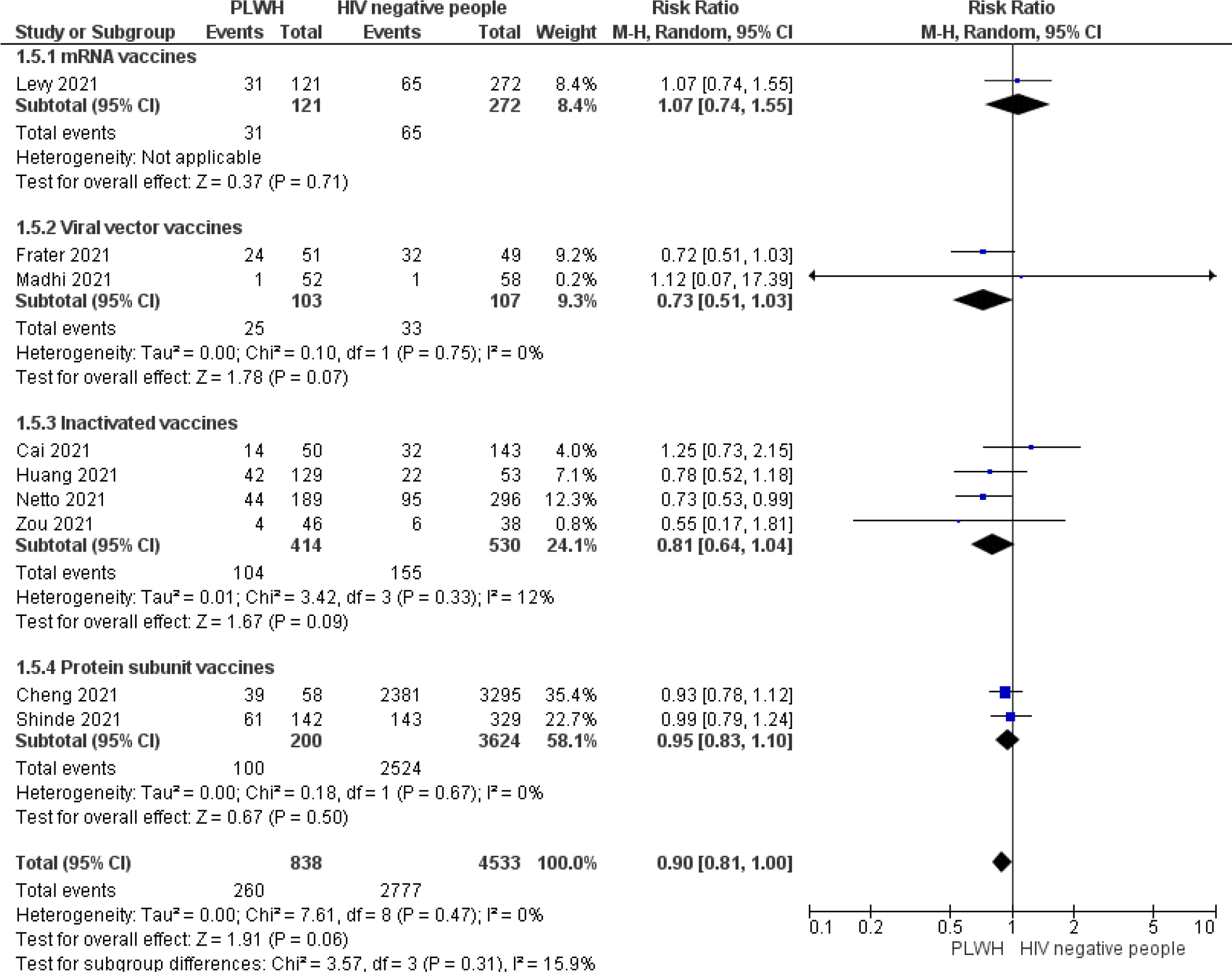
Forest plot of local reactions between PLWH and HIV negatives after 2^nd^ dose of SARS-CoV-2 vaccines

#### 3.2.11. Any systemic reaction after the 2^nd^ dose

The pooled results of systemic reactions after the 2^nd^ dose of SARS-CoV-2 vaccines were not statistically significant between 591 PLWH and 942 HIV negative people RR 0.97 (95%CI: 0.82-1.14, P = 0.71) (**Figure 13**). In contrast, PLWH had less systemic reaction after the second dose compared to HIV negative subjects’ RR 0.69(95%: 0.53- 0.91, P = 0.009) (**Figure 13**).

**Figure 13:**
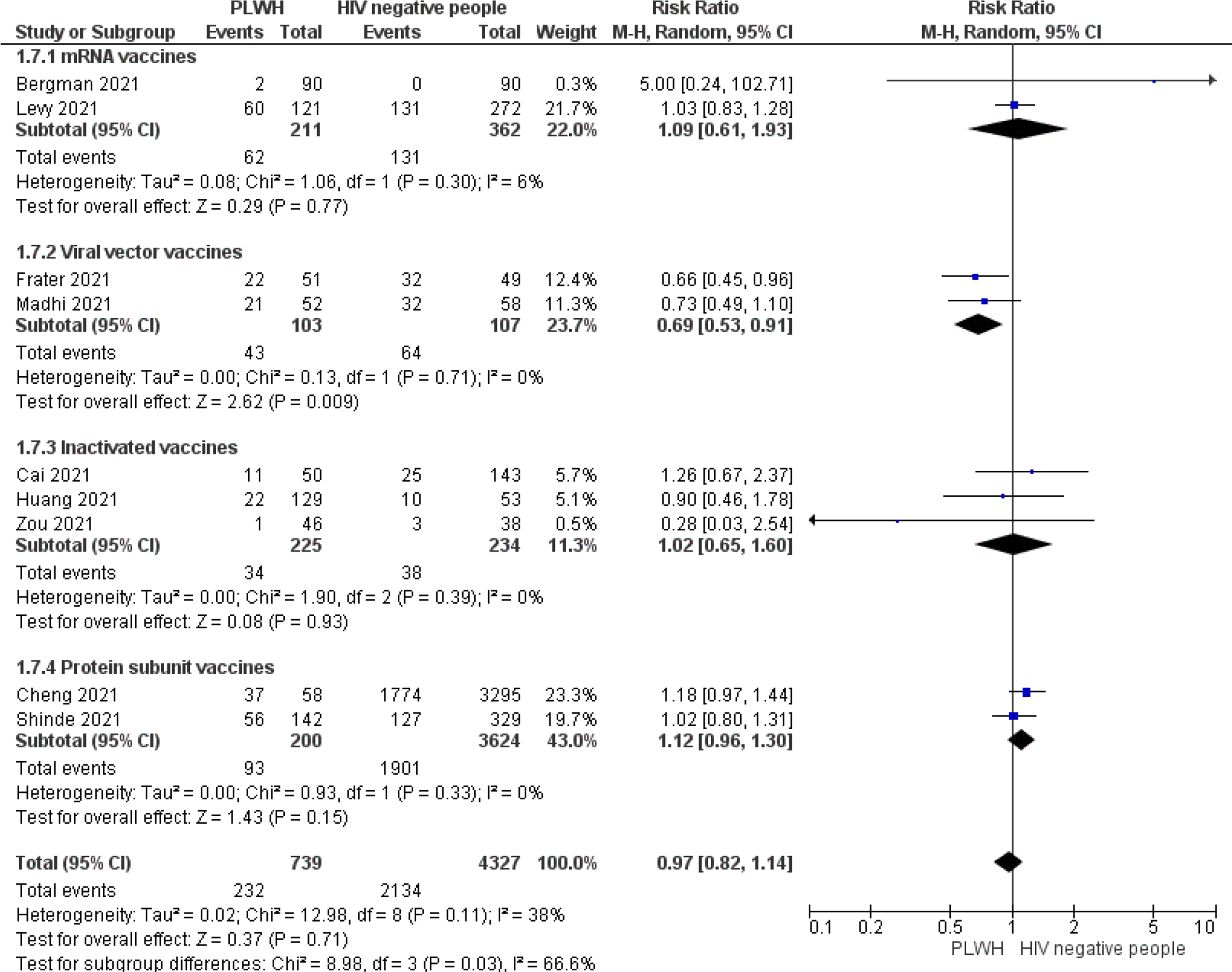
Forest plot of systemic reactions between PLWH and HIV negatives after 2nd dose SARS-CoV-2 vaccines

#### 3.2.12. IgG seroconversion based on CD4 count (<500 vs cells/µL) among PLWH ≥500

The pooled result of IgG seroconversion based on CD4 count being less and above or equal to 500 cells/ µL included 638 PLWH and five studies [47, 56–58, 61] (**Figure 14**). The result revealed no statistically significant results between the two PLWH groups RR (0.79, 95%CI: 0.63-1.00, P = 0.05) (**Figure 14**). Heterogeneity was high between the vaccine types (I^2^ = 88, P <0.001), however, the test for subgroup analysis was not statistically significant between the vaccine groups (I2 = 24.2%, P = 0.27) (**Figure 14**).

**Figure 14:**
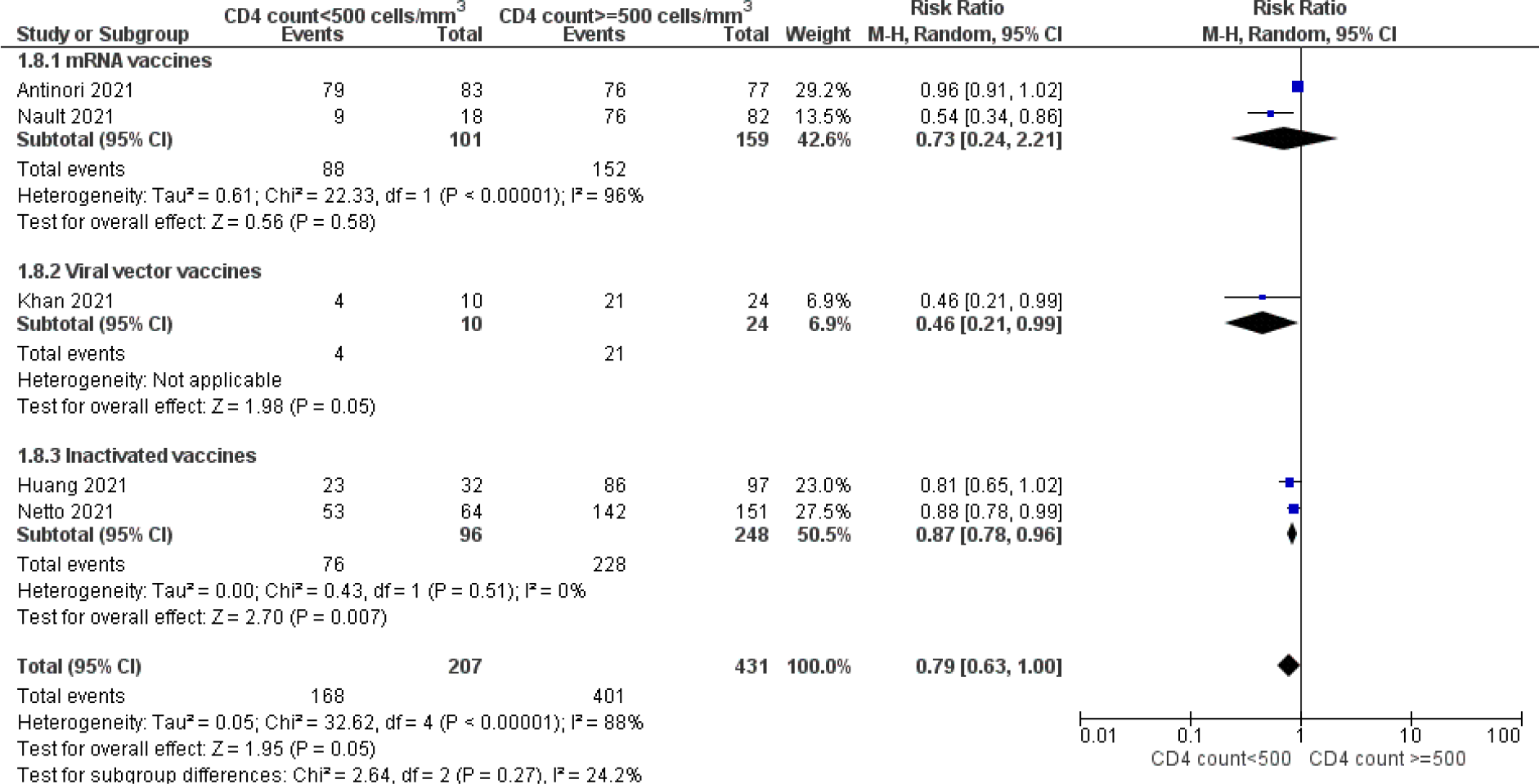
Pooled results of IgG seroconversion among PLWH based on CD4 count <500 or ≥500 cells/µL

#### 3.2.13. IgG seroconversion based on CD4 count (<200 vs cells/µL) among PLWH ≥200

**Figure 15** depicted the overall results of IgG seroconversion among 294 PLWH based on CD4 count <200 or ≥200 cells/µL. The result did not show a statistically significant difference between the two groups RR 0.68 (95%: 0.32-1.45, P = 32). The test for subgroup analysis was not statistically different between the two groups (I2 = 61, 6%, P = 0.11).

**Figure 15:**
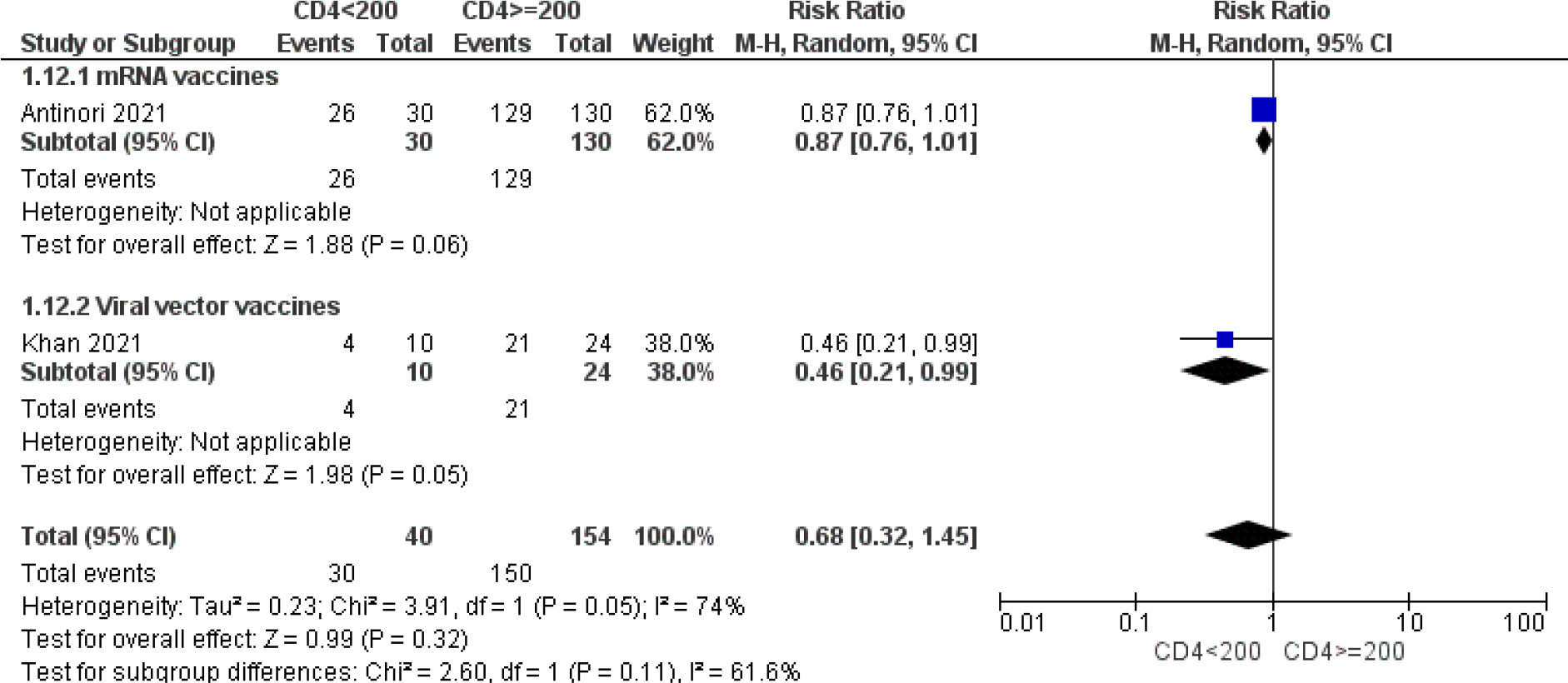
Pooled results of IgG seroconversion among PLWH based on CD4 count <200 or ≥200 cells/µL

#### 3.2.14. SARS-CoV-2 vaccines neutralizing antibodies response among PLWH (<500 vs ≥500 cells/µL)

The pooled result of neutralizing antibodies response of three studies [56, 58, 60] revealed that PLWH with CD4 less than 500 cells/µL had 15% risk reduction of neutralizing antibodies response compared to those with CD4≥500 cells/µL (RR 0.85, ≥95%CI: 0.77-0.95, P = 0.003) (**Figure 16**).

**Figure 16:**
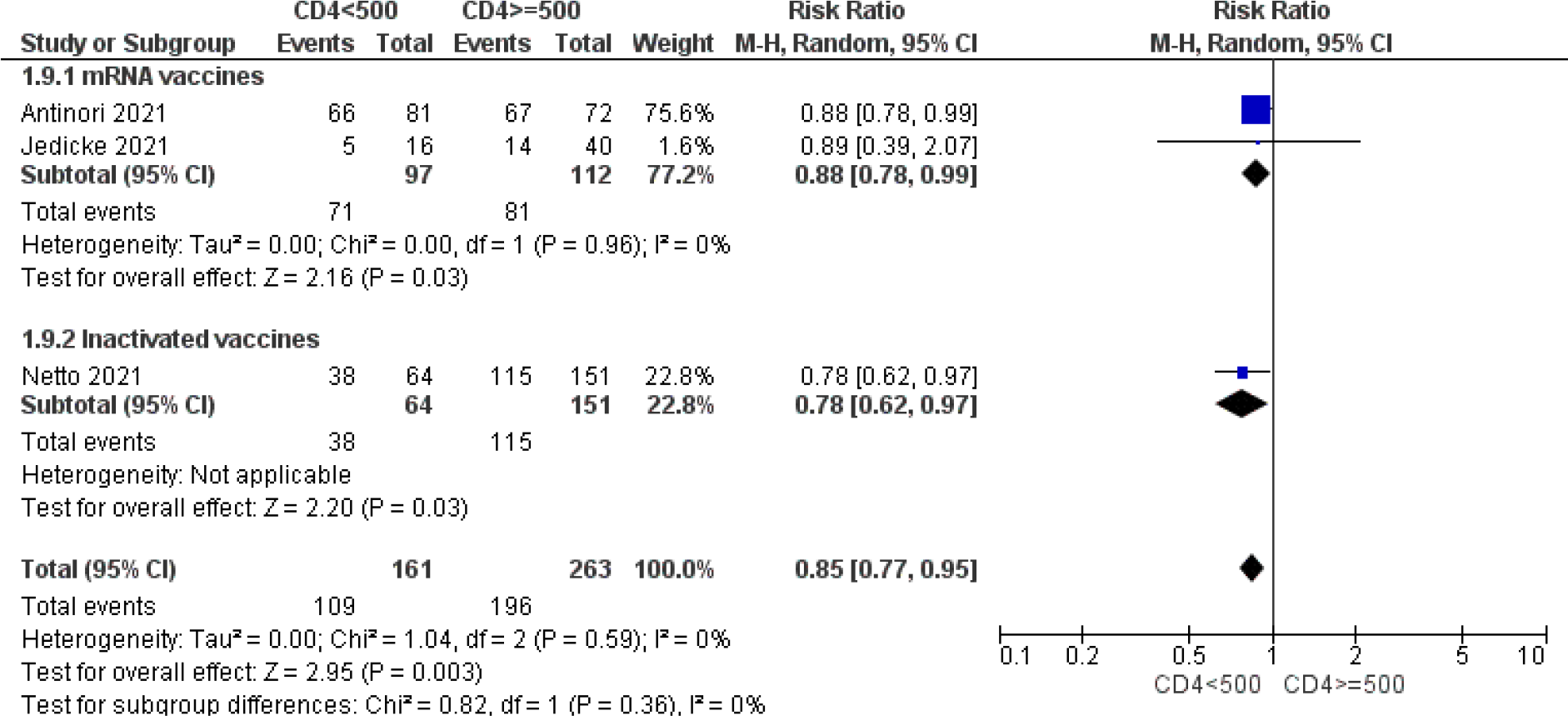
Pooled results of neutralizing antibodies response among PLWH (<500 vs ≥500 cells/µL)

### 3.3. Sensitivity analysis

To assess the robustness of our pooled RR of IgG seroconversion, IgG antibodies level, neutralizing antibodies response, vaccine efficacy, local and systemic reactions after the first and second dose of SARS-CoV-2 vaccines, we performed a sensitivity analysis. This analysis revealed that the overall estimates of IgG seroconversion, IgG antibodies level, neutralizing antibodies response, local and systemic reactions were robust and were not influenced by a single study. However, the overall estimate of vaccine efficacy was influenced by [62].

### 3.4. Meta-regression

Univariate meta-regression analyses were performed to investigate whether the median age and CD4 account were associated with the SARS-CoV-2 IgG antibodies level effect estimates. Twelve studies [16, 44, 45, 51–56, 58, 60, 61] reporting the median age of CD4 count were included. Univariate meta-regression showed than neither the median age nor CD4 cells/µl were associated with the SARS-CoV-2 IgG antibodies level (BAU/ml) effect estimates with the coefficient =0.15 and P= 0.739 (**Figure 17 a**) and coefficient = -0.00 and P = 0.910 (**Figure 17b**), respectively.

**Figure 17.**
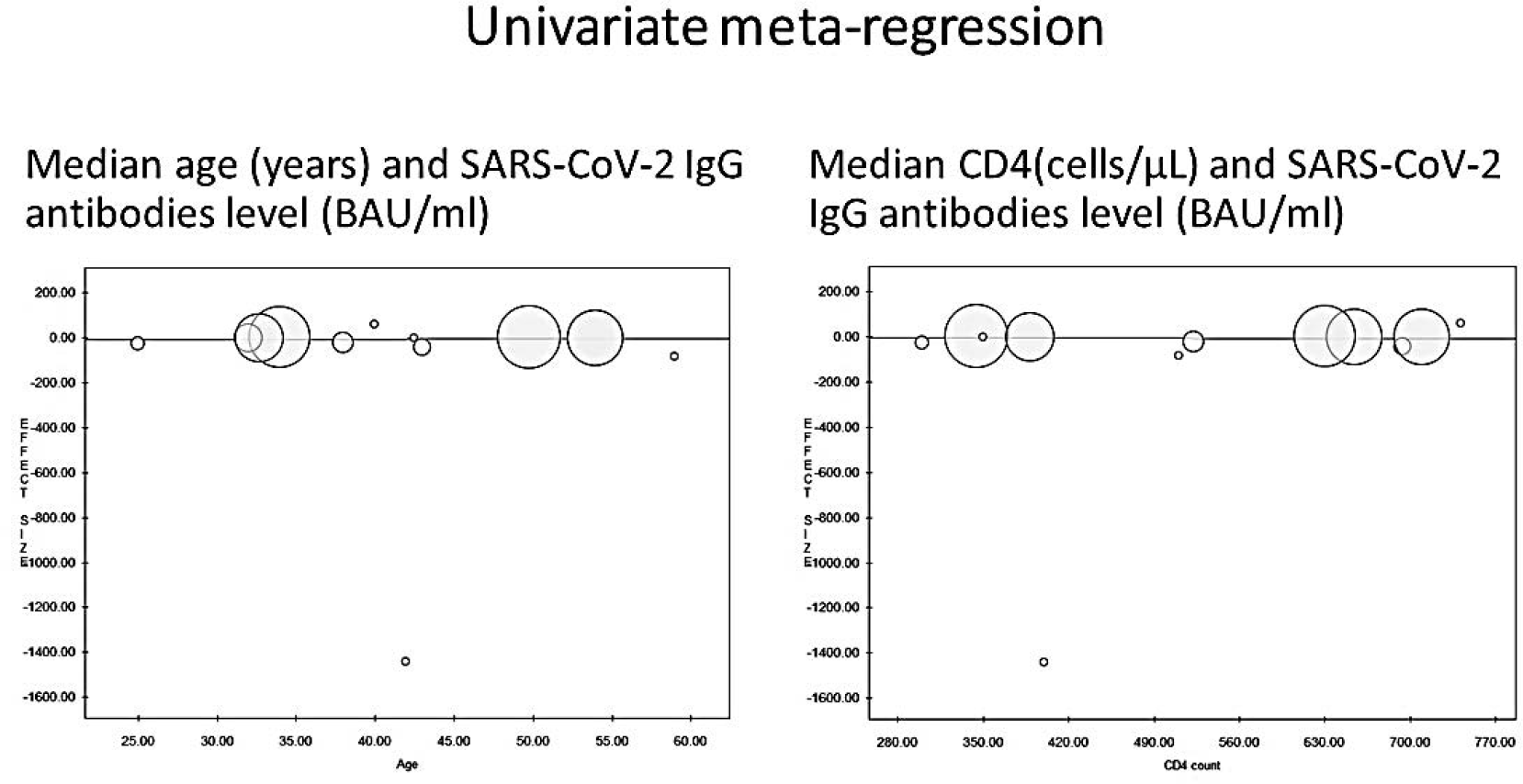
a: Meta-regression of Median age (years) and SARS-CoV-2 IgG antibodies level (BAU/ml). Figure 17 b: Median CD4(cells/µL) and SARS-CoV-2 IgG antibodies level (BAU/ml)

**Figure 18a.**
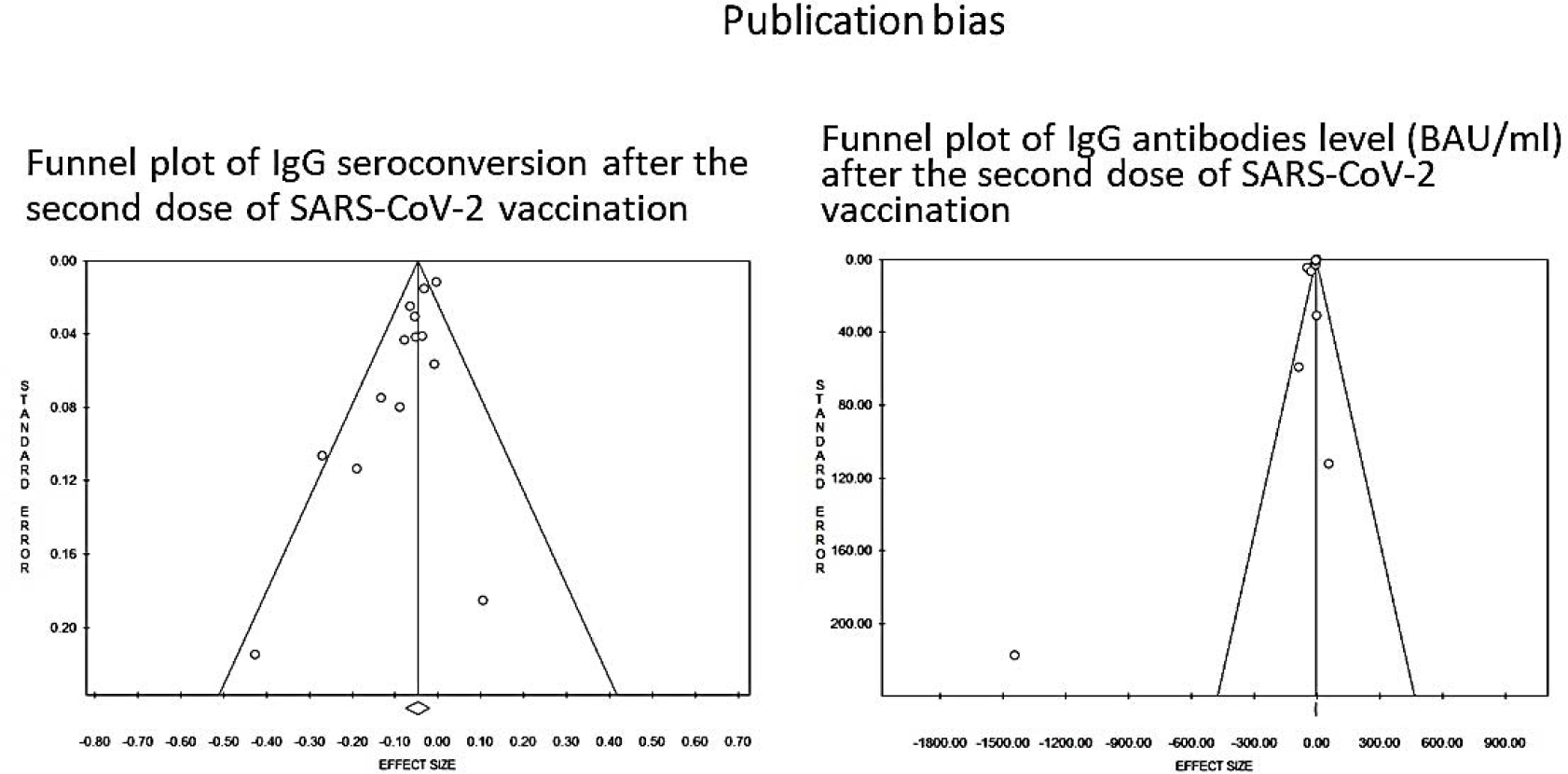
Funnel plot of IgG seroconversion after the second dose of SARS-CoV-2 vaccination. Figure 18b: Funnel plot of IgG antibodies level (BAU/ml) after the second dose of SARS-CoV-2 vaccination

### 3.5. Publication bias

As one of the pooled results included more than 10 studies, we built the funnel plot and Egger’s linear regression and Begg and Mazumdar’s rank correlation tests were used to assess publication bias. The symmetry of the funnel plots was examined to search for possible publication bias or even heterogeneity. The funnel plot for IgG seroconversion after the second dose of SARS-CoV-2 vaccines and IgG antibodies level were both asymmetrical around a single peak (Figure 16). The result of Egger’s test for small-study effects showed that there was a statistically significant publication bias in estimating IgG seroconversion after the second SARS-CoV-2 vaccination with Egger’s linear regression (t = -3.53 and P = 0.004) and Begg and Mazumdar’s rank correlation test, Z value for Kendall’s tau = -1.53, p = 0.112. In the same, the Egger’s linear regression for IgG antibodies level (t = -3.34 and P = 0.007) and Begg and Mazumdar’s rank correlation test, Z value for Kendall’s tau = -0.85, p = 0.393. This means that the weighted regression test detected statistical evidence of bias.

## 4. Discussion

This systematic review and meta-analysis assessed SARS-CoV-2 vaccines’ efficacy, safety, and tolerability in PLWH. The review included twenty-three studies and 28, 246 participants among whom 79.55% (22, 469/28, ≥246) were PLWH. The studies included 18 years. All studies included PLWH with median CD4 count cells/µL. Sixteen out of twenty-three studies reported the HIV viral load. Among them, six studies included PLWH with viral load < 50 copies/ml [16, 45, 49, 51, 53, 54] and ten others included PLWH with viral load 58.1% to 95% suppressed [44, 46, 52, 55, 56, 58, 60, 61, 63, 65].

Our findings highlighted that PLWH had lower IgG seroconversion and neutralizing antibodies response after the second vaccination dose compared to HIV negatives. However, neutralizing IgG antibodies were not different between the two groups for inactivated vaccines. The review also found that IgG antibody levels after the second dose of SARS-CoV-2 vaccines were lower in PLWH as reported in 8 out of 12 studies. However, three studies involving mRNA vaccines and heterogeneous (mRNA and viral vector vaccines) showed statistically higher MD than viral vector and inactivated vaccines [53, 58, 60]. Meta-regression revealed no relationship between the level of IgG antibodies and the median CD4 count greater than 280 cells/L or age greater than 18 years. When compared to other SARS-CoV-2 vaccines, mRNA vaccines elicited a high level of IgG antibodies [66]. By the way, mRNA vaccines produced higher levels of antibodies after the second dose than the other vaccine groups. After the first dose of SARS-CoV-2 vaccination, IgG antibody levels in PLWH and HIV-negative participants were the same. This is significant because previous research has shown that SARS-CoV-2 IgG seroconversion in PLWH was quickly lost, implying that PLWH were at a disadvantage following COVID-19 vaccination [67]. According to a recent study, data are currently insufficient to determine whether there is a significant decline in vaccine effectiveness against any form of clinical illness caused by SARS-CoV-2 infection beyond 6 months after vaccination [68]. According to Israeli data, approximately 40% of breakthrough infections occur in immunocompromised individuals [68].

Our study also revealed that IgG seroconversions between <500 vs ≥500 cells/µl or <200 vs ≥200 cells/µl were not different among PLWH. However, the neutralizing antibodies response was lower in PLWH with CD4 count <500 cells/µl than ≥500 cells/µl. This should be taken in the context of a small sample size and missing data to assess the impact of lower CD4 counts (< 200 and <500 cells/µL) on SARS-CoV-2 vaccines immunogenicity and antigenicity in PLWH. It should be noted that our included studies had the median CD4 > 200 cells/µl. Based on low immunogenicity and antigenicity among PLWH and knowing that severe immunosuppression due to HIV/AIDS is defined as a current CD4 count of <200 cells/µl for adults or children [22]. Besides, the effectiveness of the SARS-CoV-2 vaccine was 65% between vaccinated and unvaccinated PLWH. In the same, the SARS-CoV-2 among PLWH with CD4 count ≥350 cells/µl was higher than those with CD4 count < 350 cells/µl. Our results may not be generalizable to PLWH with CD4+ T-cell counts <200 cells/ µl as the sample size was very small in this specific subgroup and the vaccine effectiveness outcome was highly influenced by one study including vector viral adenovirus vaccine [62]. Besides, non-replicating vaccines, including those licensed for SARS-CoV-2 can be given to all PLWH, although replicating vaccines should be avoided in those with CD4 count < 200 cells/µl [16, 49].

Currently, CDC is recommending that moderately to severely immunocompromised people receive an additional dose [69]. The U.S Food and Drug Administration also authorized the use of an additional dose of COVID-19 vaccines in people who are immunocompromised [70, 71], reporting that a booster dose would provide enhanced protection against COVID-19 in PLWH. The new recommendations are only about a third dose to increase the level of protection from COVID-19 for PLWH with CD4 count < 200 cells/µl [72]. As a general consideration, PLWH with CD4 counts <200 cells/ µl who are not on ART should first receive ART before vaccination [73]. Vaccination should be deferred until a clinically significant immune reconstitution has been achieved, preferentially after the CD4 cell count has increased >200 cells/mm3 [73]. In fact, there are several reasons why COVID-19 vaccine booster doses may be needed in PLWH: (i) waning protection against infection or disease, in particular severe disease, over time (i.e., waning immunity), (ii) reduced protection against a variant(s) of concern (VOC), or (iii) inadequate protection from the currently recommended primary series for some risk groups for which evidence from long term SARS-CoV-2 IgG seroconversion is lacking [73]. Additionally, evidence shows PLWH with a low CD4 count, high HIV viral loads, and lack of ART use respond less strongly to vaccination, which is why they may be offered a third dose [22]. In the same line, vaccination responses for PLWH may also be suboptimal despite ART, and the duration of seroprotection was shorter in PLWH [74–76], necessitating adjusted vaccine schedules or a booster [16, 77]. However, the rationale for booster doses may differ by vaccine product, epidemiological setting, risk group, and vaccine coverage rates [73]. According to a recent study, PLWH with well-controlled viral loads on antiretroviral therapy and CD4+ T-cell counts in the healthy range may not need a third COVID-19 vaccine dose as part of their initial immunization series, though other factors such as older age, co-morbidities, vaccine regimen type, and vaccine response durability will influence when this group may benefit from additional doses [51]. Further, ART should be initiated as soon as possible, not only to preserve immune function and avoid secondary HIV transmission but also to avoid the complicated interaction between HIV infection and the above factors [78, 79]. While antibody levels may be measured, without a clear understanding of the correlates of protection against severe disease and the interaction of immune suppression with measured immune responses, clinical inferences based on the measurement of antibody levels in persons who are immunosuppressed are difficult [80]. For instance, low antibody levels may not denote poor protection against severe disease; and conversely, high antibody levels in a person unable to generate a commensurate cellular response may not denote good protection against severe disease [80]. Our review has shown high variability of anti-RBD levels between mRNA, viral vector, and inactivated SARS-CoV-2 vaccines after the second dose even though anti-RBD levels were quite constant after the first dose. This high variability can be due to the presence, in naturally infected individuals, of neutralizing antibodies other than anti-RBD (such as antibodies that recognize the N-terminal domain of the S protein) [41]. Moreover, the emergence of VOC, i.e., variant strains that carry mutations potentially able to reduce the neutralizing efficacy after either natural infection or vaccination has further increased the risk of a diminished efficacy [81]. This is important to emphasize that males were predominant in most of the included studies. However, a study reported no differences by age and gender were not significant with anti-RBD median values were evaluated in 1:20 or ≥1:80 [41]. Besides, univariate meta-regression has shown that IgG seroconversion was not correlated with the median age of 25 to 60 years and CD4 > 280 cells/µl.

Our study also showed that SARS-CoV-2 vaccines tolerability was the same between PLWH and HIV negatives in point of view local reactions after the first and second doses of SARS-CoV-2 vaccines and systemic reactions after the 2nd dose. Local reactions after the second dose of SARS-CoV-2 vaccines, on the other hand, varied significantly between studies, and the results were not consistent enough due to the small number of studies. Our findings showed that after the second dose of SARS-CoV-2 viral vector vaccine in PLWH, local reactions were significantly reduced [16, 45]. Similarly, mRNA vaccine significantly reduced systemic reactions after the first SARS-CoV-2 vaccines dose in PLWH [44], while HIV negative people had higher systemic reactions after the first SARS-CoV-2 protein subunit vaccines dose than PLWH [46]. However, those results should be considered with caution because of the small sample size among included studies. Concerns for adverse effects (AEs) significantly impact persistent vaccine hesitancy among PLWH. Studies have shown a low level of SARS-CoV-2 vaccines acceptability among PLWH compared to participants not living with HIV. In China (57.2% vs 91.3%) [82, 83], Canada (65.2% vs 79.6%) [84], India (46.1% vs 74.5) [83, 85], in French (71.3% vs 58.9%) [83, 86]. Our findings may play a substantial role to enhance COVID-19 vaccination among PLWH as fears of severe side effects are a major concern in this group population [86].

According to our knowledge, this is the first systematic review and meta-analysis to investigate the safety, tolerability, and efficacy of SARS-CoV-2 vaccines in HIV-infected patients. Little research has been conducted on this topic because PLWH are a unique population due to the severity and mortality rates of COVID-19. The review strength is the fact that the sensitivity analysis, the overall estimate of RR of IgG seroconversion, IgG antibodies levels, neutralizing antibodies response, local and systemic reactions after the first and second doses of SARS-CoV-2 vaccines was robust and was unaffected by a single study. However, the vaccine efficacy was influenced by a single large sample size study [62].

Publication was not well controlled with Egger’s linear regression test (t= -3.53 and p = 0.004) and Begg and Mazumdar’s rank correlation test (Z value for Kendall’s tau = -1.53 and p = 0.112) for IgG seroconversion after the second dose of SARS-CoV-2 vaccines and Egger’s linear regression test (t= -3.37 and p = 0.007) and Begg and Mazumdar’s rank correlation test (Z value for Kendall’s tau = 0.85 and p = 0.393) for IgG antibodies level after the second dose of SARS-CoV-2 vaccines. However, the review finding is limited by the nature of included studies. Most studies included had small sample sizes, and long-term SARS-CoV-2 IgG seroconversion, level, neutralizing antibodies, and vaccine efficacy was not assessed. Larger studies will be needed to confirm and extend the findings, particularly in the case of IgG antibodies level after the second dose of SARS-CoV-2 vaccination in PLWH. CD4 and CD8 counts should be prioritized, as SARS-CoV-2 vaccine immune responses are dependent on CD4 counts in PLWH. Further research into suppressed versus non-suppressed HIV viral load is also required. In the same line, more research on PLWH who are not receiving antiretroviral therapy and/or have low CD4+ T-cell counts is required.

Besides, further consideration is that persistent infection in severely immunocompromised PLWH (CD4 count < 200 cells/µL) may allow the emergence of variant strains of SARS-CoV-2 (variants of concerns) with the potential to escape immune responses [22]. This is critical for determining the efficacy of SARS-CoV-2 vaccines in this population. However, due to the small sample size and omission of this group in various studies, the review was unable to highlight this. Adenovirus vaccine vectors and HIV acquisition risk are also highlighted. Even though this outcome was included in the study protocol [24], we did find studies related to it. According to preliminary research, adenovirus type 5 (Ad5) immune complexes activate the dendritic cell–T cell axis, which may enhance HIV-1 replication in CD4 T cells [87, 88]. Additionally, Ad5-specific CD4 T cells may be more susceptible to HIV infection [88, 89]. A participant-level meta-analysis of six randomized trials in which 224 participants received recombinant adenovirus serotype-5 (rAd5)-vector preventive HIV-1 vaccines and 179 participants received a placebo revealed an increased risk of HIV-1 infection by the vaccine overall follow-up time, but no evidence of an increased risk of HIV-1 infection by DNA/rAd5 [90]. Due to the small sample size of most of the included trials, vaccination with a rAd5 vector increased mucosal T cell activation, which remains a central hypothesis to explain the potential enhancement of HIV acquisition in SARS-CoV-2 vaccines based on the rAd5 vector. In the future, large efficacy trials of such vaccines will be required.

Other study limitations were some data were taken from interim reports and the supplementary data provided with them. These documents had limited information in the summarized format and the individual patient data was not available, thus there is a possibility to miss some important aspects. The main limitations to this analysis are related with the intrinsic weaknesses of the included studies as shown in the risk of bias assessment (**Supplemental materials Table 2 & 3**). However, the results showed almost perfect inconsistency, indirectness, and imprecision.

## 5. Conclusion

According to our findings, PLWH had lower immunogenicity and antigenicity than HIV-negative people after the second dose of SARS-CoV-2 vaccines. However, the reactogenicity was the same after the first and second doses of SARS-CoV-2 vaccines. The antigenicity was lower among PLWH with CD4<500 cells/µl. In contrast, the immunogenicity was not different between PLWH with CD4<500 vs ≥500 cells/µl as well as CD4 count <350 vs ≥350 cells/µl. In the same line, immunogenicity among PLWH with CD4 count <200 cells/µl should be considered in the context of a small sample size.

SARS-CoV-2 vaccines tolerability after the first and second dose was not different between PLWH and HIV negatives. However, systemic reactions after the first and second doses of SARS-CoV-2 vaccines varied between studies depending on the type of vaccine. Protein subunit vaccine reported a lower risk of systemic reactions after the first SARS-CoV-2 vaccine among PLWH. In the same line, both mRNA and viral vector vaccines reported a lower risk of systemic reaction among PLWH.

Based on the above, we recommend an additional dose of SARS-CoV-2 vaccination to PLWH with CD4 count ≥200 cells/µl, low HIV viral loads, and on ART.

Our findings should be interpreted considering several study limitations, including the study’s small sample size and lack of long-term assessment of SARS-CoV-2 immunogenicity as an ideal vaccine that should provide rapid, multifaceted, long-term protection. Large-scale prospective studies, including long-term antibody monitoring, are required in the future for PLWH. Furthermore, more research should be conducted to determine whether giving a third primary dose to PLWH with a CD4 count < 200 cells/l is unlikely to result in significant harms or disadvantages but may offer the possibility of benefit.

## 6. Abbreviations

ACE2: angiotensin-converting enzyme 2; ART: Antiretroviral therapy; AU: BAU: BHIVA: British HIV Association; CD4+ T: CD8+ T: COVID-19: coronavirus disease 2019; DNA: Deoxyribonucleic acid; ELISA: enzyme-linked immunosorbent assay; FDA: Food and Drug Administration; GAVI: Global Alliance for Vaccines and Immunisation GRADE: Grading of Recommendations, Assessment, Development, and Evaluation; HAART: highly active antiretroviral therapy; HIV: Human Immunodeficiency virus; IgG immunoglobin G; mRNA: messenger ribonucleic acid; rAd5: recombinant adenovirus type 5; RBD: SARS-CoV-2 Spike Protein; ROBIN-1: PLWH: people living with HIV; PRISMA: Preferred Reporting Items for Systematic Reviews and Meta-Analyses; SARS-CoV-2: severe acute respiratory syndrome coronavirus 2; VOC: variant(s) of concern; WHO: World Health Organization

## 7. Footnotes

### Authors’ contributions

The project was initiated by JLT and PSN. JLT registered the review into Prospero and conducted an electronic search of various databases. The data extraction and risk of bias assessment were carried out by JLT and LMM in collaboration with PSN. JLT carried out a statistical analysis. JLT wrote the manuscript with the help of PSN and AM. The manuscript was reviewed and edited by JLT, PSN, LMM, and AM. The final version of the manuscript was approved by all authors.

### Availability of data and materials

All relevant data for the study are included in the article or are available as supplemental materials.

### Ethics approval and consent to participate

Not applicable.

### Consent for publication

Not applicable.

## Data Availability

All data produced in the present work are contained in the manuscript

