## Supplemental table 1 for "Systematic review and meta-analysis of COVID-19 vaccines safety, tolerability, and efficacy among HIV-infected patients"

**Supplementary material Table 1: Characteristics of included studies: systematic review and meta-analysis of COVID-19 vaccines safety, tolerability, and efficacy among HIV-infected patients**

| **Study ID/Country** | **Population/size and mean age/Baseline CD4 count and HIV VL** | **Study designs** | **Vaccine’s type/Dose/Injection process/**  **Control** | **Outcomes** | |
| --- | --- | --- | --- | --- | --- |
|  |  |  |  | Primary outcomes | Secondary outcomes |
| Levy 2021  Israel | 143 PLWH, aged >=18 years.  -Controls: 399 and 272 immunocompetent healthcare workers (-Age, years, mean (SD) range: 49.8 (11.5) 24-84  -Sex, n (%) males: 131 (92%)  -Nadir CD4+ T cell/mm3, mean (range): 345 (2-900)  -Viral load on diagnosis copies/mL, mean (range): 626, 518 (42-13M) | Prospective open study | BNT162b2 mRNA vaccination (the first and the second vaccines, respectively). | **IgG seroconversion**  After 1^st^ dose  HIV+: 66/128  HIV-: 66/128  After 2^nd^ dose  HIV+: 139/141  HIV-: 269/272  **IgG Antibody response level (BAU/ml)** (Mean ± SD)/n  After the 1^st^ dose  HIV+: 0.18(0.4)/128  HIV-: 0.18(0.4)/128  After the 2^nd^ dose  HIV+: 5.20(0.51)/141  HIV-: 6(0.43)/272 | **Local reactions**  After 1^st^ dose  HIV+: 54/133  HIV-: 102/399  After 2^nd^ dose  HIV+: 31/121  HIV-: 65/272  **Systemic reactions**  After 1^st^ dose  HIV+: 26 /133  HIV-: 192/399  After 2^nd^ dose  HIV+: 60/121  HIV-:131/272 |
| Frater 2021  UK | 54 adults with HIV, aged 18-55 years (median age 42.5 years (IQR 37.2-49.8),  All participants were on ART, suppressed HIV viral loads and CD4+ T cell counts >350 cells/µl at enrolment.  Sex (male): 54 (100.0%) | open-label non-randomised | A prime-boost regimen of ChAdOx1 nCoV-19, with two doses (5 × 1010 vp) was given 4-6 weeks apart. | **IgG seroconversion**  After 1^st^ dose  HIV+: 40/44  HIV-: 25/31  After 2^nd^ dose  HIV+: 29/43  HIV-: 17/28  **Antibody response level (BAU/ml)**  **(Mean±SD)/n**  HIV+: 164.84(132.83)/49  HIV-: 165.92(131.96)/28 | **Local reactions**  After 1^st^ dose  HIV+: 40/53  HIV-: 44/50  After 2^nd^ dose  HIV+: 24/51  HIV-: 32/49  **Systemic reactions**  After 1^st^ dose  HIV+: 40/53  HIV-: 43/50  After 2^nd^ dose  HIV+: 22/51  HIV-: 32/49 |
| Madhi 2021  South Africa | 52 PLWH received the vaccine and 58 controls received either the vaccine or a placebo. Median (IQR) age 37 (32–45) (HIV group), and 32 (25–42) (control group)**;** Median (IQR) CD4+ count, cells/μL: 742 (540–953) and Median (IQR) detectable VL, copies/mL: 30 (10–105); Median age: 40(33-46) and Male%: 27/103 | Phase 1b/2, double-blind, randomized, placebo-controlled trial. | PLWH and HIV-negative participants were randomized 1:1 to receive two intramuscular injections of ChAdOx1 nCoV-19 or saline placebo (0.9% sodium chloride), separated by an interval of 28 days. | **IgG seroconversion**  After 1^st^ dose  HIV+: 25/36  HIV-: 15/23  After 2^nd^ dose  HIV+: 28/32  HIV-: 22/23  **IgG Antibody response level (BAU/ml)**  (Mean±SD)/n  After the first dose  HIV+: 134.11(180.45)/36  HIV-: 93.71(144.09)/23  After the second dose  HIV+: 502.57(524.18)/32  HIV-: 443.23(307.48)/23  **SARS-CoV-2 post vaccination**  HIV+: 8/104  HIV-: 24/70 | **Local reactions**  HIV+: 1/52  HIV-:1/58  **Systemic reactions**  HIV+: 21/52  HIV-: 32/58 |
| Shinde 2021  South Africa | 240 participants ≥18 to <65 years of age in Cohort 2 were HIV-positive. All participants with HIV were medically stable, taking HIV treatment and with a viral load below 1000 copies/ml. | Randomized control trial | Participants were randomly assigned in a 1:1 ratio to receive two intramuscular injections, 21 days apart, of either NVX-CoV2373 (5 μg (subunit) of recombinant spike protein with 50 μg of Matrix-M1 adjuvant) or saline placebo (injection volume, 0.5 ml) | **IgG seroconversion**  HIV+: 46/109  HIV-: 1368/2684  **SARS-CoV-2 post vaccination**  HIV (+vac): 4/109  HIV+ (plac): 2/102 | **Local reactions**  After 1^st^ dose  HIV+: 70/150  HIV-:137/334  After 2^nd^ dose  HIV+: 61/142  HIV-: 143 /329  **Systemic reactions**  HIV+: 56/142  HIV- :127/329 |
| Haidar 2021  USA | 37 HIV-infected people. Female 3 (8.1%) and male 34 (91.9%) | Prospective cohort study | Most participants (98.2%) received the mRNA-1273 (Moderna=48.5%) or BNT162b2 (Pfizer=49.7%) vaccine series | **IgG seroconversion**  After 2nd dose  HIV+: 35/37  HIV-: 105/107  **Antibody response level (BAU/ml)**  (Mean±SD)/n  After 2^nd^ dose  HIV+: 14.04(16.83)/37  HIV-: 16.22(16.25)/107 |  |
| Janssen 2021  USA | 2.7% (n=601) had stable/well-controlled HIV received Ad26.COV2. S  2.8% (n=617) had HIV received placebo; CD4 cell count ≥300 cells/μL within 6 months prior to screening HIV viral load <50 copies/mL within 6 months. | Randomized control trial, in the phase 3 trials | Ad26.COV2. S and placebo in the subgroup of HIV positive participants | **IgG seroconversion**  After 1^st^ dose  HIV+: 323/467  HIV-: 361/498  After 2^nd^ dose  HIV+:316/461  HIV-:356/493  **SARS-CoV-2 post vaccination**  HIV+: 2/461  HIV+ (Plac): 4/493 |  |
| Moderna 2021  USA | 176 people living with HIV out of 30,000 participants, in adults 18 years of age and older. | A Phase 3,  randomized,  stratified,  observer-blind,  placebo controlled study | The Moderna COVID-19 Vaccine, mRNA-1273 (100 μg) is administered intramuscularly as a series of two doses (0.5 mL each), given 28 days apart. | **Serology against SARS-CoV-2 nucleocapsid**  HIV+(vac): 80/80  HIV+(plac): 75/76  **SARS-CoV-2 post vaccination**  HIV+(vac): 1/90  HIV+(plac): 1/86 |  |
| Nault 2021  Canada | HIV-positive individuals (n=121) were recruited, stratified according to their CD4 counts (<250, 250-500 and above 500 cells/µL. A control group of 20 health care workers naïve to SARS CoV-2 was used. Mean [range] age was 43 [21, 65] years. Males: 90 (84.9%) | Prospective cohort study | mRNA vaccine | **Positive RBD IgG**  HIV+: 100/106  HIV-: 19/20 |  |
| Brumme 2021  Canada | A total of 100 adult PLWH and 152 control participants without HIV. All PLWH were receiving antiretroviral therapy; the most recent plasma viral load <50 [<50 - <50] The CD4+ T-cell count was 710 (IQR 525-935; range 130-1800) cells/µL. Age in years, median [IQR] 54 [40-61]. | Prospective cohort study | mRNA vaccine  ChAdOx1  ChAdOx1/mRNA | **IgG Antibody response level (BAU/ml)**  (Mean±SD)/n  After 1^st^ dose  HIV+:0.23(0.08)/66  HIV-: 0.27(0.08)/148  After 2^nd^ dose  HIV+:34.42(7.54)/96  HIV-: 36.65(6.9)/151 |  |
| Zou 2021  China | 46 PLWH and 38 HNC.  All PLWH were on ART, 89% had virus suppressed (41/46), and  the median CD4 count of PLWH was 523 cells/μL  The mean (SD) age of PLWH was 38 (9) years  and 87% were males. | Prospective cohort study | Inactivated COVID-19 vaccine at day 0 and the second dose at day 28. | **IgG seroconversion**  After 1^st^ dose  HIV+: 12/46  HIV-: 24/38  After 2^nd^ dose  HIV+: 19/46  HIV-: 24/38  **IgG Antibody response level (BAU/ml)**  (Mean±SD)/n  After 1^st^ dose  HIV+: 4.46 (1.92)/46  HIV-: 18.28 (26.71)/38  After 2^nd^ dose  HIV+: 4.58(1.92)/46  HIV-: 26.13(26.71)/38  **NAb positivity**  HIV+: 26%/46  HIV-: 63%/38 | **Local reactions**  After 1^st^ dose  HIV+: 14/46  HIV-: 11/38  After 2^nd^ dose  Local  HIV+: 4/46  HIV-: 6/38  **Systemic reactions**  After 1^st^ dose  HIV+: 5/46  HIV-: 2/38  After 2^nd^ dose  HIV+: 1/46  HIV-: 3/38 |
| Ogbe 2021  UK | PWH (N=54; all male), Participants had undetectable VL 156 (<50 HIV RNA copies/ml) and a median CD4 count of 694 cells/µl (IQR 573.5 –859.5) | open-label, non-randomised sub-study | mRNA – mRNA  ChAdOx1 – mRNA  ChAdOx1- ChAdOx1 | **Antibody response (BAU/ml)**  (Mean±SD)/n  After 1^st^ dose  HIV+: 0.23(0.08)/54  HIV-: 0.23(0.08)/60  After 2^nd^ dose  HIV+: 105.26(7.44)/54  HIV-: 147.73(38.65)/60 |  |
| Feng 2021  China | Healthy individuals (n=28) and HIV-1 infected on (cART) (n=42). CD4+ T cell count of above 200 cells/μL with VL below and beyond 20 copies/ml. | open-label two-arm non-randomized study | inactivated COVID-19 vaccine | **NAb positivity**  After 1^st^ dose  HIV+: 25%/42  HIV-: 10%/28  After 2^nd^ dose  HIV+: 32/42  HIV-: 21/28  **Antibody response(BAU/ml)**  **(Mean ± SD)/n**  After 1^st^ dose  HIV+: 0.29(0.11)/42  HIV-: 0.29(0.11)/28  After 2^nd^ dose  HIV+: 24.54(6.50)/42  HIV-: 29(5.93)/28 |  |
| Spinelli 2021  USA | 100 HIV infected adults and 100 HIV-negative patients. Nearly 90% were men and the median age was 59 years. The median CD4 was 511 and five people had detectable viral load (HIV RNA above 200 copies). CD4: 511 (351-796). | matched case-control observational study | Two doses of the Pfizer or Moderna vaccines. Three quarters received the Pfizer vaccine, and a quarter received the Moderna vaccine. | **IgG seroconversion**  HIV+:88/100  HIV-:95/100  **Antibody response level (BAU/ml)**  **(Mean±SD)/n**  After 2^nd^ dose  HIV+: 205.73(330.25)  HIV-: 291.27(493.12)  **NAb positivity**  HIV+: 76/100  HIV-: 88/100 |  |
| Netto 2021  Brazil | 511 participants (215 PLWH and 296 controls). CD4+ T cell counts were <500 cells/mm3 for 64 (30%) participants and ≥500 cells/mm3 for the remaining 151 (70%). Overall, 191 (89%) PLWH had undetectable (<50 copies/mL) viral load. Age: Median Age (IQR): 54 (45-60). 48 (37 – 58) | Prospective controlled study | SARS-CoV-2 inactivated vaccine CoronaVac | **Positive RBD IgG**  After 1^st^ dose  HIV+: 41/214  HIV-: 114 / 295  After 2^nd^ dose  HIV+: 185 / 204  HIV-: 265 / 274  **Antibody response level (BAU/ml)**  **(Mean±SD)/n**  After 1^st^ dose  HIV+: 0.78(1.19)/215  HIV-: 4.89(14.6)/296  After 2^nd^ dose  HIV+: 10.21(11.17)  HIV-: 12.71(8.25)  **NAb positivity**  After 1^st^ dose  HIV+:40/211  HIV-:112 / 289  After 2^nd^ dose  HIV+:143 / 202  HIV-: 229 / 274 | **Local reactions**  After 1^st^ dose  HIV+:23/189  HIV-:71/296  After 2^nd^ dose  HIV+: 44/189  HIV-: 95 /296  CD4 <500 cells/mm3  Positive RBD-binding  IgG (%): 53/64  NAb positivity: 38/64  CD4≥500  Positive RBD-binding  IgG (%): 142/151  NAb positivity: 115/151 |
| Khan 2021  South Africa | 26 PLWH and 73 HIV-uninfected participants. CD4 count cells/μL: 735 and the mean age was 45 years | Prospective cohort study | Ad26.CoV2. S vaccine | **IgG seroconversion**  After 2^nd^ dose  HIV+: 23/26  HIV-: 73/73 | CD4<200  Positive RBD-binding: 4/10  CD4>500  Positive RBD-binding: 21/24 |
| Antinori 2021  Italy | 166 PLWH (SID: CD4 count<200/mm3=32; MID (CD4 count: 200-500/mm3) =56; NID (CD4 count >500/mm3) =78) on ART. Proportion of PLWH with HIV-RNA lower than 50 copies/mL was 68.8% in SID, 92.9% in MID, and 100% in NID and 71.6% were female with a median age of 42 years. | Observational cohort study | BNT162b2 or mRNA-1273 vaccines. As vaccination, 93 (57%) received BNT162b2, and 70 (43%) mRNA-1273. | **IgG seroconversion**  After 2^nd^ dose  HIV+: 155/160  HIV-: 168/168  **Antibody response level (BAU/ml) (Mean±SD)/n**  After 2^nd^ dose  HIV+:1563.83(1250.53)/160  HIV-: 3008.8(2512.01)/168  **NAb response**  HIV+: 133/153  HIV-: 72/73 | **CD4 <500**  Positive RBD-binding  IgG (%):79/83  NAb positivity: 66/81  **CD4:>=500**  Positive RBD-binding  IgG (%):**76/77**  NAb positivity: 67/72 |
| Cai 2021  China | A total of 143 PLWHs (PLWH group) and 50 healthy controls (control group) were included. The mean (SE) age was 31.56 ± 8.05 years and the mean CD4 was 398.96 cells/µL. | Non-interventional cross-sectional study | SARS-CoV-2 inactivated vaccine | **IgG seroconversion**  After 2^nd^ dose  HIV+: 83/143  HIV-: 38/50  **RBD-IgG** GMT (Mean±SD)/n  After 2^nd^ dose  HIV+: 1.58 ± 1.19/143  HIV-: 2.33 ± 1.65/50  **NAb response**  HIV+:65/67  HIV-:19/20  **NAb response**  (Mean±SD)/n  HIV+: 27.8 ± 18.69  HIV-: 19.82 ± 22.95 | **Local reactions**  After 2^nd^ dose  HIV+: 14/50  HIV-: 32/143  **Systemic reactions**  After 2^nd^ dose  HIV+: 11/50  HIV-: 25/143 |
| Jedicke 2021  Germany | All PLWH had a viral load of ≤ 200 HIV-1 RNA copies/mL and 96.5% had a viral load of < 50 copies/mL.  CD4 first dose: 716 (151–1558), CD4 second dose: 577 (45–1106) | Prospective cohort study | BNT162b2 (mRNA) vaccination | **Antibody response level (BAU/ml)**  **(Mean±SD)/n**  After 1^st^ dose  HIV+: 13.52(26.50)/56  HIV-: 20.38(11.33)/41  After 2^nd^ dose  HIV+: 45.54(48.05)/50 HIV-: 71.24(13.67)/41  **NAb response**  HIV+:47/52  HIV-:41/41 | **CD4 counts below and above 500 cells/µL**  CD4 <500  median anti-S IgG  26.23 RU/mL; n = 16; interquartile range (IQR) 148.1 RU/mL  CD4:>=500  vs. 33.35 RU/mL; n = 40; IQR 86.78 RU/mL |
| Huang 2021  China | 129 PLWH and 53 HIV-negative individuals with median (IQR) age of 34 (28, 38). Over half of them had an undetectable viral load (58·1%), and the median CD4+ T-cell count was 630·5 (IQR: 499·5, 848·8). | A cross-sectional study | A least one dose of inactivated SARS-CoV-2 vaccine. Participants received Sinovac-CoronaVac (55·0% versus 30·2%, P<0·001) and only completed the prime dose (27·1% versus 3·8%) | **IgG seroconversion**  HIV+: 120/129  HIV-: 52/53  **Antibody response level (BAU/ml)**  **(Mean±SD)/n**  HIV+: 1.23(1.02)/129  HIV-: 2.22(2.35)/53  **Neutralizing antibody**  HIV+: 92/129  HIV-: 52/53 | **Local reactions**  HIV+: 42/129  HIV-: 22/53  **Systemic reactions**  HIV+:22 /129  HIV-: 10/53  **Neutralizing antibody levels.**  CD4<500:23/32  CD4>=500: 86/97 |
| Vladimir 2021  Russia | All were PLWH, vaccinated (n=2543), unvaccinated (n=17592) and uncompleted vaccination (n=4288) with CD4 count of 639[484 - 821], 526[334 - 730] and 586[424 - 774], respectively. Mean age  (M±SD): 44·69±10·12, 41·50±10·11 and 41·65±8·28 | Retrospective cohort study | Sputnik V (Gam-COVID-Vac) vaccine | **SARS-CoV-2 post vaccination**  HIV+vacc: 71/2543  HIV+unvac: 1354/16606 | **SARS-CoV-2 infection**  Vaccinated  CD4+ < 350: 11/231  CD4+ >=350: 51/ 1967  Unvaccinated  CD4+ < 350: 352/ 2997  CD4+ >=350: 779/ 8352 |
| Zhengchao 2021  China | Twenty-four HIV-positive individuals and 24 healthy donors. 50% were male among PLWH. Age, [Median (Q3- Q1), years]: 44.00 (39.00,48.75). Absolute CD4 count: >300 cells/µL | Clinical trial | Twenty-four HIV-positive individuals who have received two injections of inactivated SARS-CoV-2 vaccines (CoronaVac, 3 μg/0.5 mL; or BBIBP-CorV, 4 μg/0.5 mL) | **Neutralizing antibodies** against SARS-CoV-2 Spike RBD  HIV+: 19/24  HIV-: 21/24 |  |
| Bergman 2021  Sweden | 90 PLWH and 90 control group. PLWH with the age <65 years, n (%):71 (79%). Latest CD4 T cell count 300 cells/ul (n= 30) and Latest CD4 T cell count >300 cells/ul (n= 60) | Open-label, non-randomized prospective clinical trial | The participants were given injections of BNT162b2 mRNA vaccine in standard dose (30 micrograms) into the deltoid muscle of the non-dominant arm on days 0 and 21 of the study; i.e., in a two-dose regimen according to the label. | **Seroconversion**  **HIV+: 89/90**  **HIV-: 90/90** | Systematic reactions  HIV+: 2/90  HIV-: 0/90 |
| Cheng 2021  Taiwan | A total of 57 PWH of ≥ 20 years of age who are on stable antiretroviral therapy and with CD4+ T cell ≥ 350 cells/mm3 and HIV viral load < 103 copies/ml were compared with 3295 HIV-negative participants. | Phase 2, prospective, double-blinded and multi-centre study | Two doses of MVC-COV1901 which is a subunit vaccine. two standard doses of 15 mcg MVC-COV1901, administered 28 days apart via IM injection. | **Seroconversion**  **HIV+: 58**/58  **HIV-: 3289/3295**  **SARS-CoV-2 neutralizing**  **GMT’s (95% CI)**  HIV+: 143.48 IU/mL (95% CI 115.5-178.3)  HIV-: and 439.01 IU/mL (95% CI 419.5-459.5) | **Local reactions**  HIV+: 39/58  HIV-: 2381/3295  **Systemic reactions**  HIV+: 37/58  HIV-: 1774/3295 |
