## Supplemental table 2 for "Systematic review and meta-analysis of COVID-19 vaccines safety, tolerability, and efficacy among HIV-infected patients"

**Supplementary material Table 2: Risk of bias assessment for randomized control trials**

| N | Study ID | Random sequence generation (Selection bias) | Allocation concealment (Selection bias) | Blinding of participants and personnel (Performance bias) | Blinding of outcomes assessment (Detection bias) | Incomplete outcome data (Attrition bias) | Selective reporting (Publication bias) | Other bias |
| --- | --- | --- | --- | --- | --- | --- | --- | --- |
| 1 | Frater 2021 | High | Unclear | Unclear | Low | Low | Low | High |
| 2 | Madhi 2021 | Low | Unclear | Low | Low | Low | Low | High |
| 3 | Shinde 2021 | Low | Unclear | Low | Low | Low | Low | High |
| 4 | Janssen 2021 | Low | Unclear | Low | Low | Low | Low | Low |
| 5 | Moderna 2021 | Low | Unclear | Low | Low | Low | Low | Low |
| 6 | Bergman 2021 | High | Unclear | Unclear | Low | Low | Low | High |
| 7 | Cheng 2021 | High | Unclear | Low | Low | Low | Low | High |
| 8 | Zhengchao 2021 | High | Unclear | Unclear | High | Low | Low | High |
