## Supplemental table 3 for "Systematic review and meta-analysis of COVID-19 vaccines safety, tolerability, and efficacy among HIV-infected patients"

**Supplementary material Table 3: risk of bias assessment for nonrandomized studies**

| N | Study ID | Domain 1: confounding | Domain 2: selection | Domain 3: classification of intervention | Domain 4: deviation from interventions | Domain 5: missing data | Domain 6: measurement of outcomes | Domain 7: selection of reported result | ROBINS-I overall | |
| --- | --- | --- | --- | --- | --- | --- | --- | --- | --- | --- |
| 1 | Levy 2021 | 1–2 | 1–2 | 1–2 | 1–2 | 2 | 1–2 | 2 | 1–2 | Low-Moderate |
| 2 | Haidar 2021 | 1–4 | 3–4 | 1 | 1–4 | 2–3 | 2–3 | 2 | 3–4 | Serious–Critical |
| 3 | Nault 2021 | 4 | 3–4 | 1 | 1–2 | 2–3 | 3 | 2 | 4 | Critical |
| 4 | Brumme 2021 | 1-2 | 2 | 1 | 1–2 | 2 | 1–2 | 1-2 | 3–4 | Low–Moderate |
| 5 | Zou 2021 | 2 | 2 | 1 | 1–2 | 2 | 1–2 | 2 | 3–4 | Low–Moderate |
| 6 | Spinelli 2021 | 2 | 1–2 | 1–2 | 1–2 | 2 | 1–2 | 2 | 1–2 | Low-Moderate |
| 7 | Netto 2021 | 2–3 | 1–3 | 1–2 | 1–3 | 3 | 2–3 | 2 | 1–3 | Low–Serious |
| 8 | Khan 2021 | 1–2 | 2–3 | 1–2 | 2 | 2 | 2 | 1–2 | 1–2 | Low-Moderate |
| 9 | Antinori 2021 | 1–2 | 1–2 | 1 | 1–2 | 2 | 2 | 2 | 1-2 | Low-Moderate |
| 10 | Cai 2021 | 4 | 3–4 | 1 | 1–2 | 2–3 | 3 | 2 | 4 | Critical |
| 11 | Jedicke 2021 | 2–3 | 1–2 | 1–2 | 2–3 | 3 | 2–3 | 2 | 2–3 | Moderate-Serious |
| 12 | Huang 2021 | 2-3 | 2 | 2 | 2–3 | 2–3 | 3 | 2–3 | 3–4 | Moderate-Serious |
| 13 | Vladimir 2021 | 1–3 | 1–3 | 1-3 | 1–2 | 2 | 2–3 | 2 | 1–3 | Low–Serious |
| 14 | Feng 2021 | 1-3 | 1 | 1 | 2-3 | 2-3 | 1-3 | 2 | 1-3 | Low-Serious |
| 15 | Ogbe 2021 | 1–2 | 1–2 | 1 | 1–3 | 2 | 1–2 | 2 | 1–3 | Low–Serious |
